# Overnutrition is a risk factor for iron deficiency in children and young people: a systematic review and meta-analysis of micronutrient deficiencies and the double burden of malnutrition

**DOI:** 10.1101/2024.01.22.24301603

**Authors:** Xiaomian Tan, Pui Yee Tan, Yun Yun Gong, J. Bernadette Moore

**Affiliations:** School of Food Sciences and Nutrition, University of Leeds, Leeds, UK

**Keywords:** Iron, zinc, vitamin A, malnutrition, obesity, systematic review and meta-analysis, children, adolescence

## Abstract

**Introduction:** Traditionally associated with undernutrition, increasing evidence suggests micronutrient deficiencies can co-exist with overnutrition. Therefore, this work aimed to systematically review the associations between iron, zinc and vitamin A status and weight status (both under- and overweight) in children and young people.

**Methods:** Ovid Medline, Ovid Embase, Scopus, and Cochrane databases were systematically searched for observational studies assessing micronutrient status (blood, serum, or plasma levels of iron, zinc, or vitamin A biomarkers) and weight status (body mass index or other anthropometric measurement) in humans under 25 years of any ethnicity and gender. Risk of bias assessment was conducted using the American Dietetic Association Quality Criteria Checklist. Where possible, random effects restricted maximum likelihood (REML) meta-analyses were performed. PROSPERO (CRD42020221523).

**Results:** After screening, 83 observational studies involving 190,443 participants from 44 countries were identified, with many studies having reported on more than one micronutrient and/or weight status indicator. Iron was the most investigated micronutrient, with 46, 28, and 27 studies reporting data for iron, zinc, and vitamin A status, respectively. Synthesizing 16 records of odds ratio (OR) from 7 eligible studies, overnutrition (overweight and obesity) increased odds of iron deficiency (OR [95%CI]: 1.51 [1.20, 1.82], p<0.0001, I^2^=40.7%). Odds appeared to be higher for children living with obesity (1.88 [1.33, 2.43], p<0.0001 I^2^=20.6%) in comparison to those with overweight (1.31 [0.98, 1.64], p<0.0001 I^2^=40.5%), although between group differences were not significant (p=0.08).

**Conclusions:** Overnutrition is associated with increased risk of iron deficiency, but not zinc or vitamin A deficiencies, with an inverted U-shaped relationship observed between iron status and bodyweight. Our results highlight significant heterogeneity in the reporting of micronutrient biomarkers and how deficiencies were defined. Inflammation status was rarely adequately accounted for, and the burden of iron deficiency may well be under-recognised, particularly in children and young people living with overnutrition.

**Key questions:** *What is already known on this topic:* -summarise the state of scientific knowledge on this subject before you did your study and why this study needed to be done

- Low-and middle-income countries are increasingly facing a double burden of malnutrition; that is, the coexistence of undernutrition (stunting, wasting, underweight) with overnutrition (overweight and obesity).
- While the relationship between undernutrition and critical micronutrients for childhood growth and development (e.g., iron, zinc, and vitamin A) is well established, less is known about the risk of micronutrient deficiencies in children and adolescents with overweight or obese, a hidden form of malnutrition.
- There are limited data summarising associations between biomarkers of the most commonly limiting micronutrients and body weight status, particularly in children and young people.

*What this study adds:* -summarise what we now know as a result of this study that we did not know before

- Overnutrition increases the risk of iron deficiency, but not zinc or vitamin A deficiencies.
- There is an inverted U-shaped relationship observed between iron status and bodyweight in children and young people, with iron deficiencies observed more frequent in both under- and overnutrition.
- Studies done to date have been heterogeneous in terms of populations studied, diagnostic criteria, and approaches to data analysis; few followed current guidelines for measuring inflammation and defining micronutrient deficiencies.

*How this study might affect research, practice or policy:* -summarise the implications of this study

- Increased recognition by healthcare practitioners that children and young people living with overweight, or obesity are likely to be iron deficient; thereby improving clinical practice and care.
- The gaps in evidence highlighted are addressed, with more research from currently underrepresented counties examining micronutrient deficiencies and the double burden of malnutrition.
- The weaknesses in study design and reporting highlighted are addressed, with improved quality and reporting of micronutrient assessment in children and young people.

## INTRODUCTION

Deficiencies in micronutrients contribute to impaired immune function, poor growth and physical development, and increased morbidity and mortality in children^1 2^. Public health prevention strategies such as supplementation, fortification, and nutrition education are therefore strongly encouraged by the World Health Organisation and UNICEF in low- and middle-income countries (LMICs)^3 4^. Although great strides have been made in the last century to address micronutrient deficiencies (MNDs) and reduce childhood mortality rates, disappointingly in 2013 (median year of data collection), the global prevalence of deficiency in one or more micronutrients was estimated to be 56% in children under 5 years old, although rates vary across countries^5 6^.

Among the micronutrients, deficiencies in iron, zinc and vitamin A remain particularly prevalent and causally associated with adverse health outcomes for children. Iron deficiency (ID) and iron deficiency anaemia (IDA) are major global health challenges affecting more than 1.2 billion people worldwide^7^. In addition to iron deficiency, it is estimated that more than 25% of South Asia and Sub-Saharan African populations are at risk of insufficient dietary zinc intake, with some countries particularly deficient^8^. In a large population survey in Ethiopia for example, the prevalence of zinc deficiency among young children was found to be a shocking 89%, improving somewhat in school children to 71%^9^. Similarly in Southeast Asia in Vietnam, in 2010 the prevalence of zinc deficiency was remarkably high (68%) in young children, improving to 42% in children over 5 years of age^10^. Moreover, although large-scale vitamin A supplementation programmes have been implemented in many countries^2^, subclinical vitamin A insufficiency is still prevalent in Vietnam (10.1%, young children aged 5-75 months old) from Vietnam nationwide food consumption survey^10^ and Africa (Ethiopia, 45.4% for 6–72 months old children from meta-analysis in 2020^11^), and often observed in the context of multiple micronutrient deficiencies^12^.

Historically MNDs were considered as one of the four forms of undernutrition, alongside wasting, stunting and underweight^13^, and a particular concern for LMICs where undernutrition may be the leading cause of childhood mortality for children under five^2 14^. However increasingly, it is recognised that MNDs also occur in the context of overweight and obesity^15^. Deficiencies in iron^16^, zinc^17^, and vitamin A^18^ have been observed in adults living with overweight and obesity and associated metabolic diseases. Typically associated with nutrient-poor, energy-dense diets, the presence of multiple micronutrient deficiencies in the absence of an energy-deficit diet has been described as ‘hidden hunger’^19^. While in high-income countries obesity is associated with ultra-processed foods that are high in fat, sugar, salt, and energy; in LMICs overweight and obesity is often associated with poverty and monotonous diets with limited choices of low-cost energy-dense staples such as corn, wheat, rice, and potatoes^19 20^. Even in high income countries, childhood obesity is strongly linked to poverty and socioeconomic and health disparities^21^.

Moreover, many LMICs are now facing a double burden of malnutrition with the co-existence of an increasing prevalence of overnutrition alongside undernutrition^22^. This is intricately linked to the rapid increase in the global prevalence of obesity, especially in 5–19 years old children in recent decades^23^. Deficiencies in multiple micronutrients including iron^24 25^, zinc^17 26^, and vitamin A^27 28^ have also been observed in children living with malnutrition. Indeed, the term ‘triple burden of malnutrition’ aims to underscore the coexistence of micronutrient deficiencies alongside undernutrition (stunting, wasting, underweight) and overnutrition^29^.

While increasing evidence now suggests micronutrient deficiencies are associated with overnutrition as well as undernutrition, to date there have been only a limited number of reports summarising associations between biomarkers of micronutrient status and obesity in children and young people. In one meta-analysis of multiple co-morbidities associated with obesity in children, higher odds ratio (OR) for both vitamin D deficiency [OR (95% CI): 1.9 (1.4, 2.5)] and iron deficiency (OR 2.1, 95% CI 1.4, 3.2) were found in children under age ten living with obesity^30^. Similarly, a separate meta-analysis of vitamin D deficiency in children and adolescents aged 0-18 years old, also found a positive association between obesity and vitamin D deficiency (OR 1.4, 95% CI 1.3, 1.6)^31^. However, to our knowledge the risks of iron, zinc, and vitamin A deficiencies, the most frequently limiting micronutrients in children and young people, have not been collectively examined. Therefore, in this work we aimed to systematically review the associations between iron, zinc and vitamin A status and body weight in children and young people, using meta-analyses to summarise where sufficient data existed.

## METHODS

This review was conducted following the Preferred Reporting Items for Systematic Reviews and Meta-Analyses (PRISMA) guidelines^32^, and was registered at Prospero (CRD42020221523).

### Search strategy

The Ovid Medline, Ovid Embase, Scopus, and Cochrane databases were systematically searched through 19 April 2023. A combination of keywords and related MeSH terms were used from three main themes: i) population: infants, children, adolescents and young adults; ii) malnutrition indicators, including under- and overnutrition; and iii) blood micronutrient indicators for iron, zinc, and vitamin A. The specific search strategies developed for each database are reported in **Supplementary Tables S1-4**.

### Inclusion and exclusion criteria

Observational studies in humans under 25 years old^33^, of any nationality, gender or ethnicity, with under- or overnutrition diagnosed by body mass index (BMI) or other anthropometric measurement, were included. Studies had to have measured as primary outcomes either: a blood or serum test of micronutrient level, OR or relative risk of micronutrient deficiencies, or the linear regression between a micronutrient indicator and weight status to be included. Records excluded from this review were: non-English articles, short reports, communications, address, case reports, comments, letters, editorial matters, meta-analyses, news, reviews, conference abstracts, studies with unrepresentative samples (either convenience sampling or n<100), studies not reporting primary outcomes of interest, studies reporting associations with combined micronutrient deficiencies, and studies of micronutrient supplement interventions. Studies that assessed anaemia diagnosed with only haemoglobin were excluded, as not specific to iron deficiency.

### Study selection

The screening of identified studies was managed using the web based Rayyan software^34^. The selection of the eligible studies for inclusion in this review was assessed and agreed by two independent reviewers (XT and PYT). Disagreement between reviewers were resolved by discussion, or by third reviewer if necessary.

### Data extraction

A standardised data extraction form was utilised to extract the following information: first author, year of publication, year of study, study design, country, participants’ characteristics (e.g., sample size, recruitment, gender, age, socioeconomic status etc.), exposure (e.g., anthropometric indicators and blood/serum levels of micronutrients of interest), control or non-exposure (e.g., children who were not malnourished), outcome measures (e.g., changes in anthropometric indicators in the presence of micronutrient deficiencies or otherwise changes in the blood micronutrients levels in different anthropometry groups), main findings (e.g., differences in mean, β coefficient, OR, relative risk), and funding or sponsorship.

### Risk of bias assessment

Risk of bias was assessed by two reviewers using the Quality Criteria Checklist developed by the Academy of Nutrition and Dietetics^35^. Any disagreement was resolved by discussion, and by a third reviewer if necessary. This tool has 10 validity questions: 1) clear research question, 2) non-biased participant recruitment, 3) group comparability, 4) report of withdrawals/participant ratio, 5) blinding, 6) study procedures description, 7) outcome, 8) appropriate statistical method and adequate adjustments, 9) conclusion supported by results, and 10) conflict of interest. The overall rating was defined as either positive (majority of criteria above were met, in which criteria 2,3,6,7 must be met), neutral (any of criteria 2,3,6,7 was not met), or negative (six or more of the criteria not being met).

### Data analysis

The associations (e.g., OR, relative risk, linear regression) between the blood micronutrient levels of interests and anthropometry status were curated in tables according to micronutrient. Mean micronutrient levels between anthropometry groups were included if there was no further analysis of data. For the convenience of summarising, the main findings of each study were concluded as either direct associations (micronutrient levels increased with increasing weight status indicators) or inverse associations (decreased micronutrient levels — increasing risk of micronutrient deficiencies— with increasing weight status indicators).

Weight status indicators included BMI-for-age z score (BAZ) and BMI used to categorise overnutrition. While for undernutrition, height-for-age z score (HAZ), weight-for-age z score (WAZ), and weight-for-height z score (WHZ) were used to categorise stunting, wasting and underweight respectively, following the WHO growth chart, obesity work taskforce, or national standard in respect to each country. Micronutrient status indicators included: blood micronutrient levels (iron, zinc, retinol), haemoglobin (Hb), iron profile, β-carotene, retinol binding protein, or diagnosis of ID, IDA, zinc deficiency or VAD. The diagnostic criteria of micronutrient deficiencies mostly followed either WHO criteria, or the International Zinc Nutrition Consultative Group standard, or country-specific standards for defining micronutrient deficiencies.

Where there were at least 5 studies^36^ assessing the risk of MNDs in malnutrition groups comparing with normal weight group, random effects restricted maximum likelihood (REML) meta-analyses were conducted to estimate the OR with 95% CI using STATA 18 (Stata Corporation, College Station, TX, US). Heterogeneity across the studies was evaluated using I^2^. Sensitivity analysis was conducted by using the leave-one-out method, to compare the pooled OR before and after eliminating each study at a time. To detect publication bias, the asymmetry of the funnel plot was examined. Statistical significance was set at P<0.05. To avoid overestimation of power, the gender-stratified data were not included in meta-analysis if overall population data were reported by the study. All graphics were produced in either STATA or using the R-package ggplot2^37^, ggalluvial^38^ and rworldmap^39^ in the R environment^40^.

## RESULTS

Utilising systematic search strategies, a total of 9711 articles were initially identified from four databases, of which, 5459 articles remained after deduplication. After title and abstract screening, the full texts of 151 articles were assessed to check for eligibility. From these, 83 observational studies met the inclusion criteria, and 7 studies were eligible for meta-analysis (**Figure 1**).

**Figure 1.**
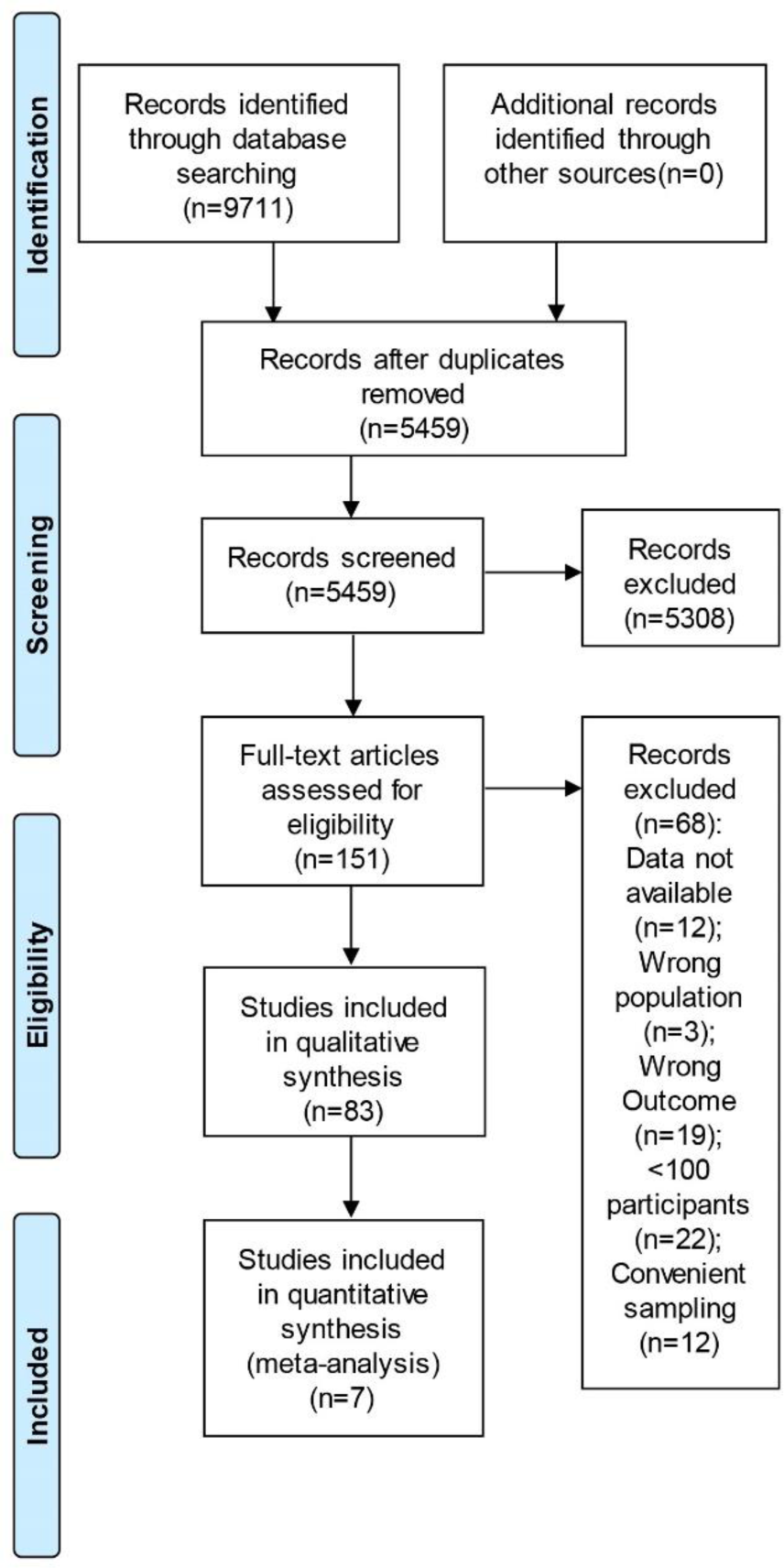
Flow diagram of the systematic identification and selection of articles.

Overall, the 83 studies involved a total of 190,443 participants from 44 countries (**Figure 2**). The majority (n=74, 89.1%) were cross-sectional studies, and the number of participants in individual studies ranged from n=101^41^ to n=64,850 (the China National Nutrition and Health Survey of Children and Lactating Mothers study, 2016–2017^42^), with a median n=675 participants. Notably, many studies assessed more than one micronutrient and/or weight status indicator at the same time. Iron was the most investigated micronutrient, with a total of 46 (n=10, 21 and 15 for both, overnutrition and undernutrition, respectively) 28 (n=4, 8 and 16 for both, overnutrition and undernutrition, respectively) and 27 (n=5, 12, 10 for both, overnutrition and undernutrition, respectively) studies identified for iron (**Table 1**), zinc (**Table 2**), and vitamin A (**Table 3**), respectively.

**Figure 2.**
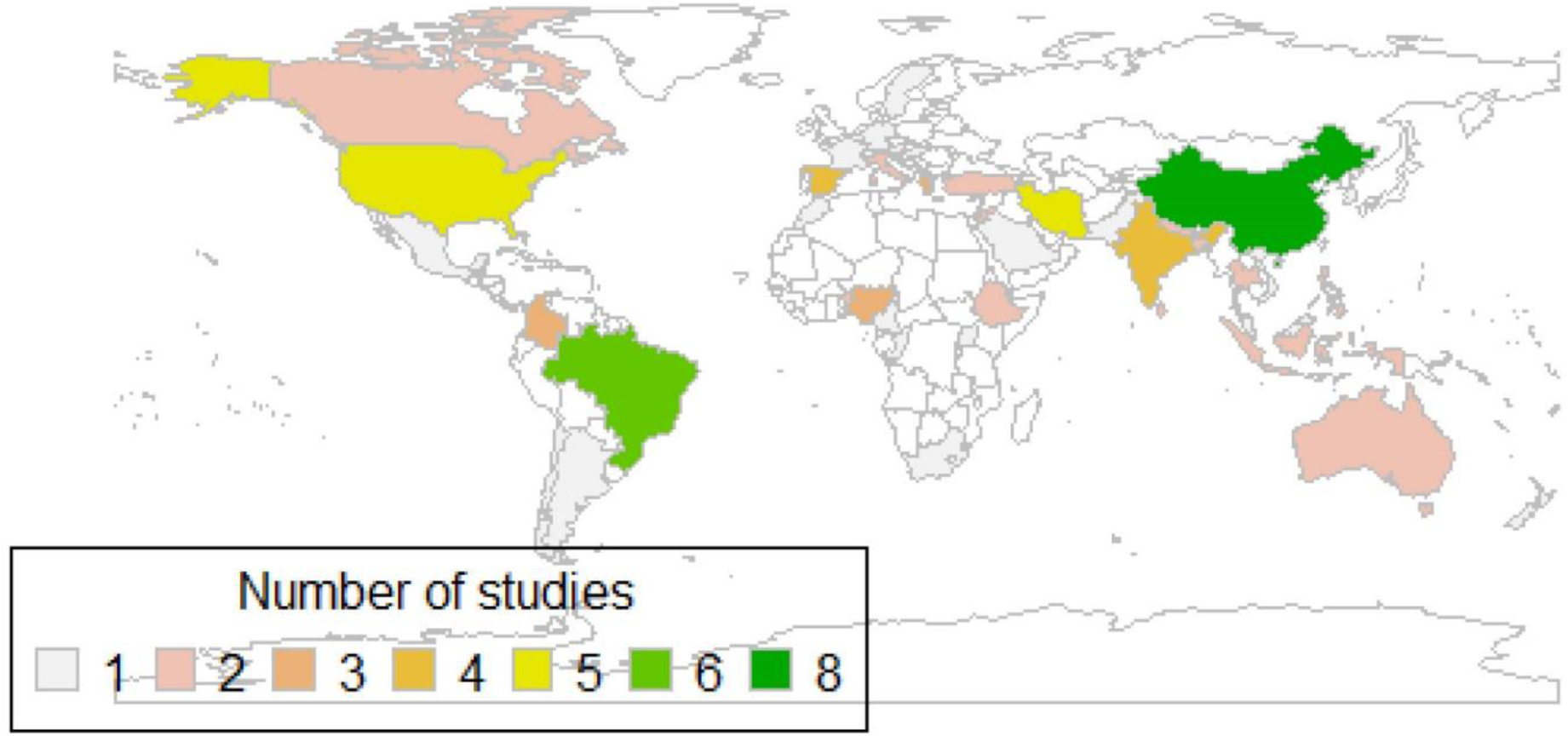
World map showing the populations represented in the included articles.

**Table 1.**
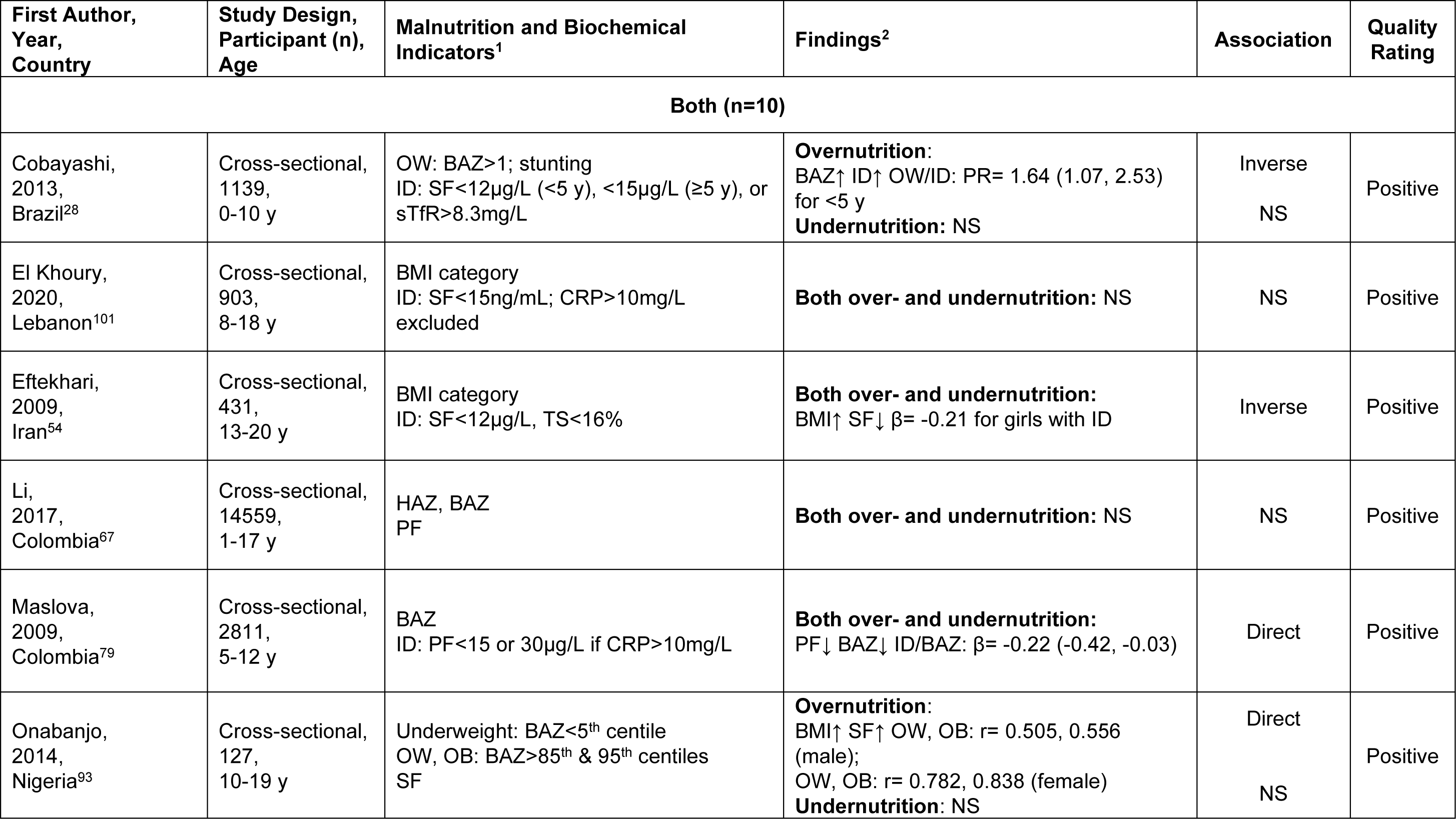
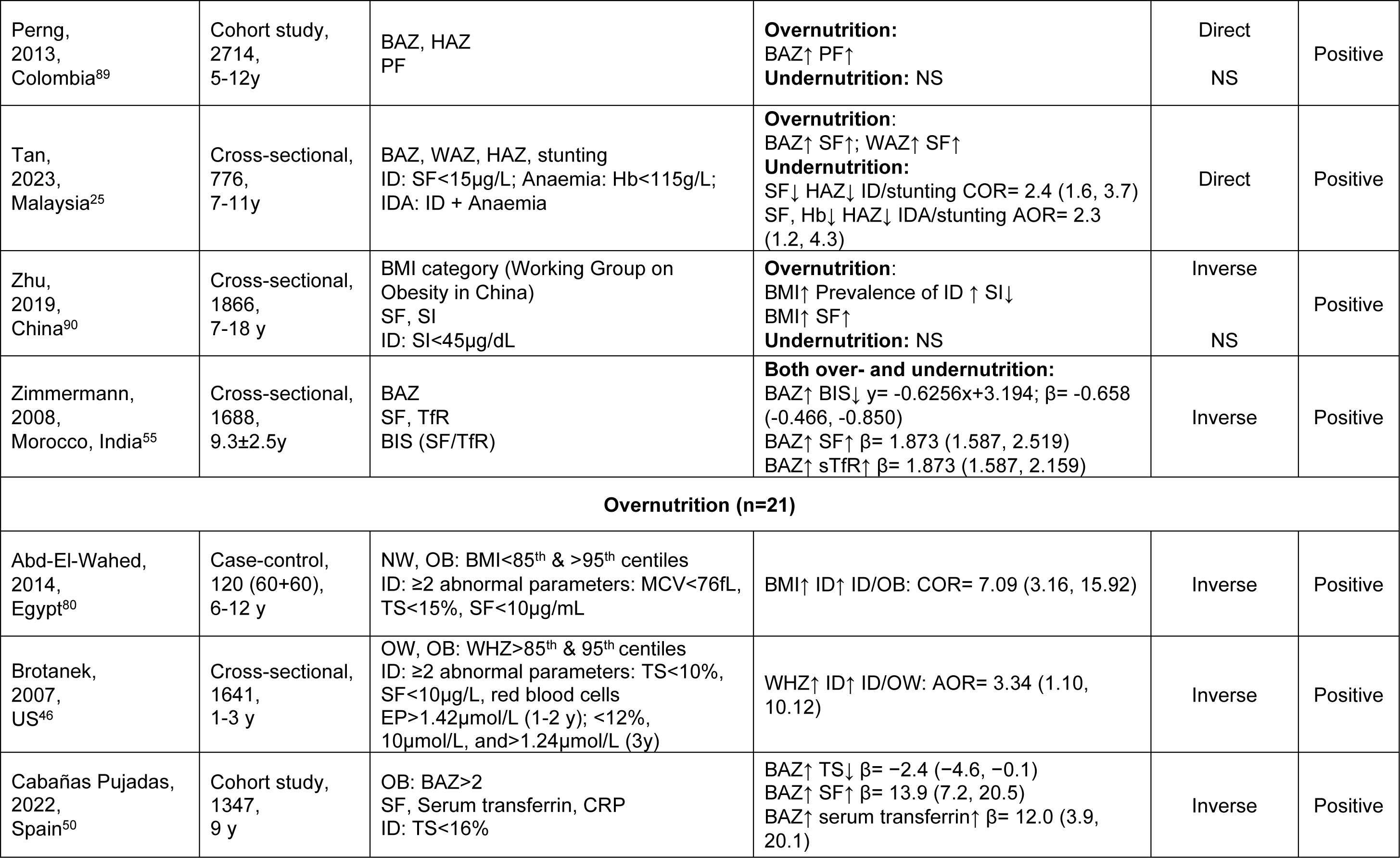
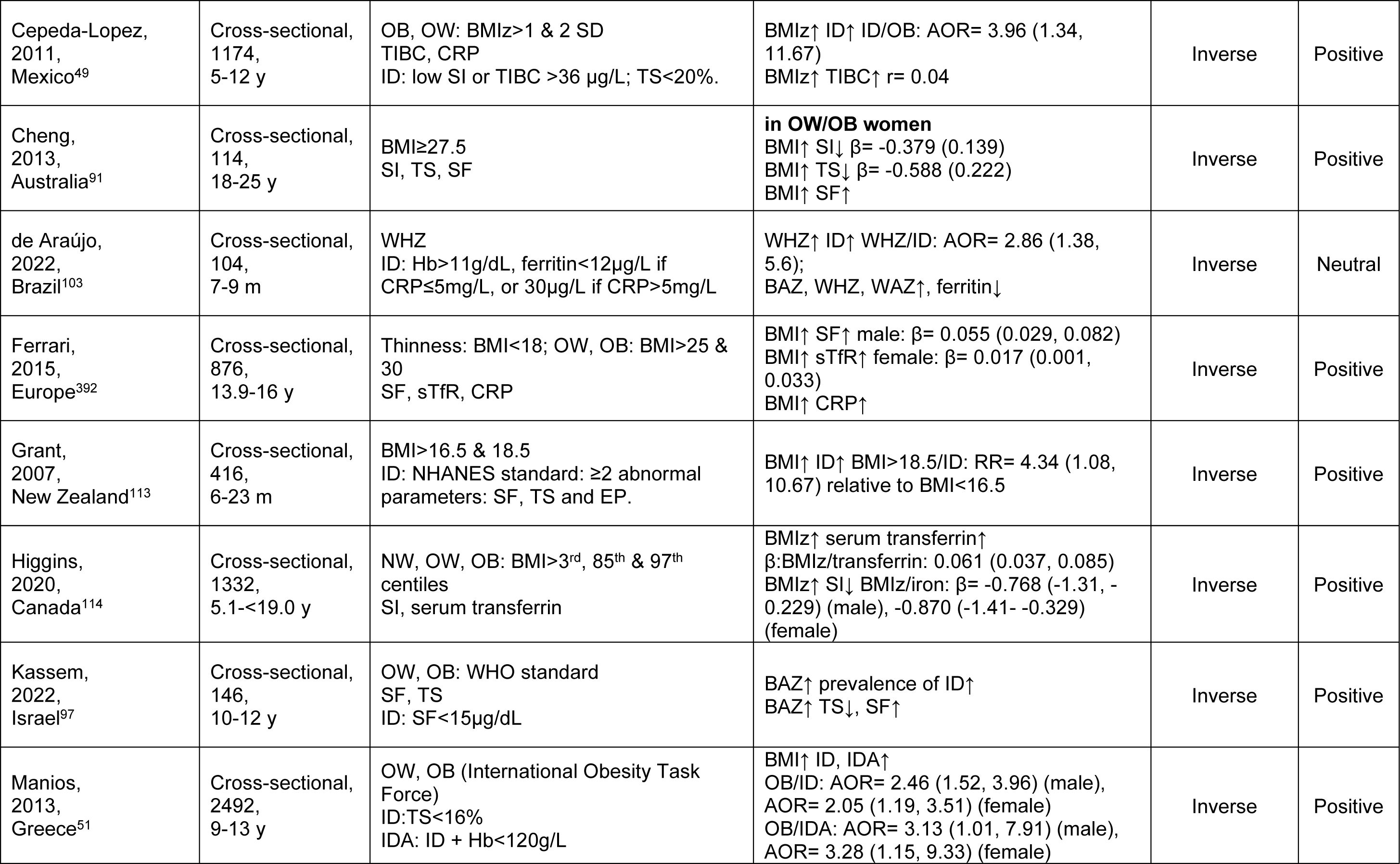
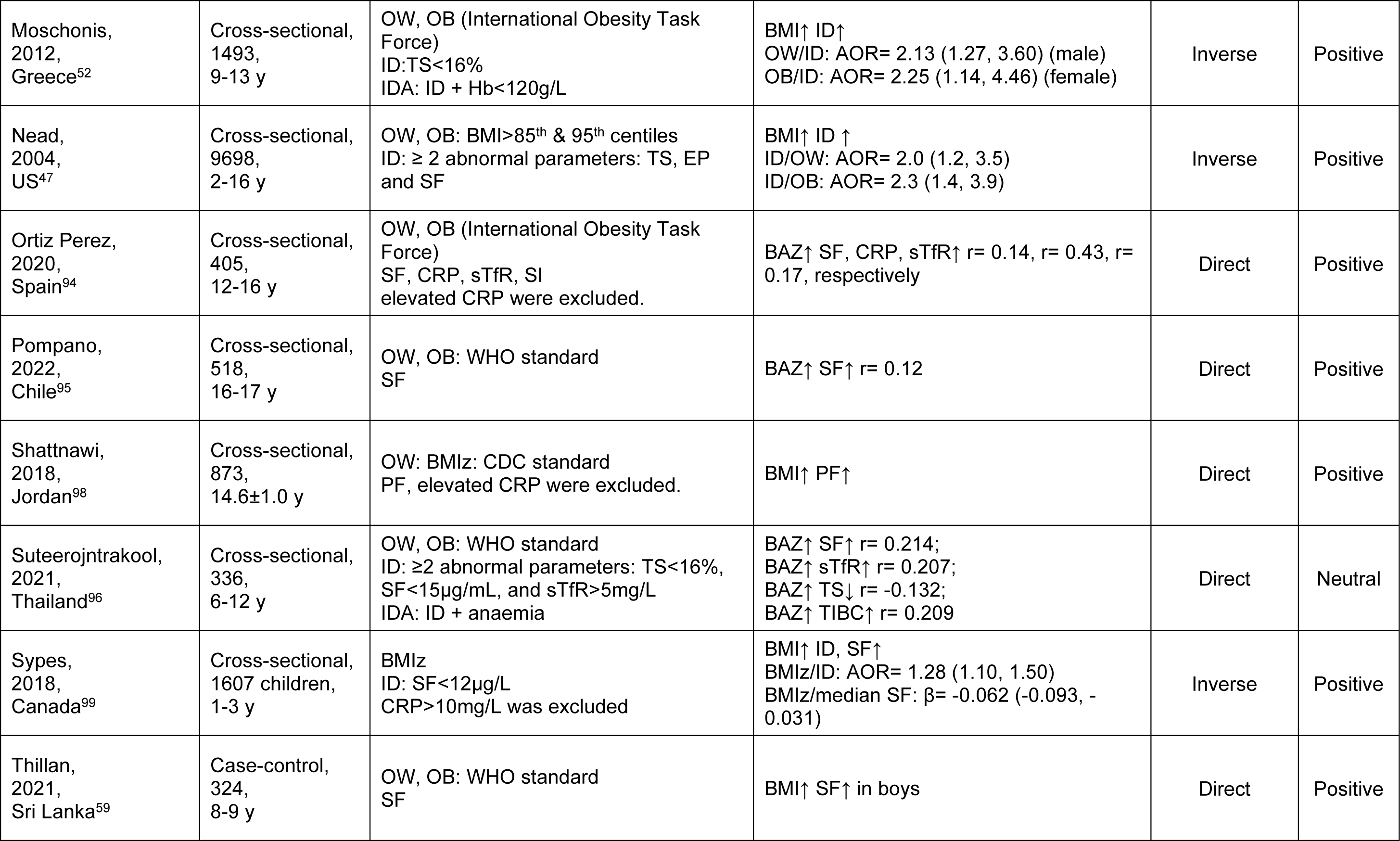
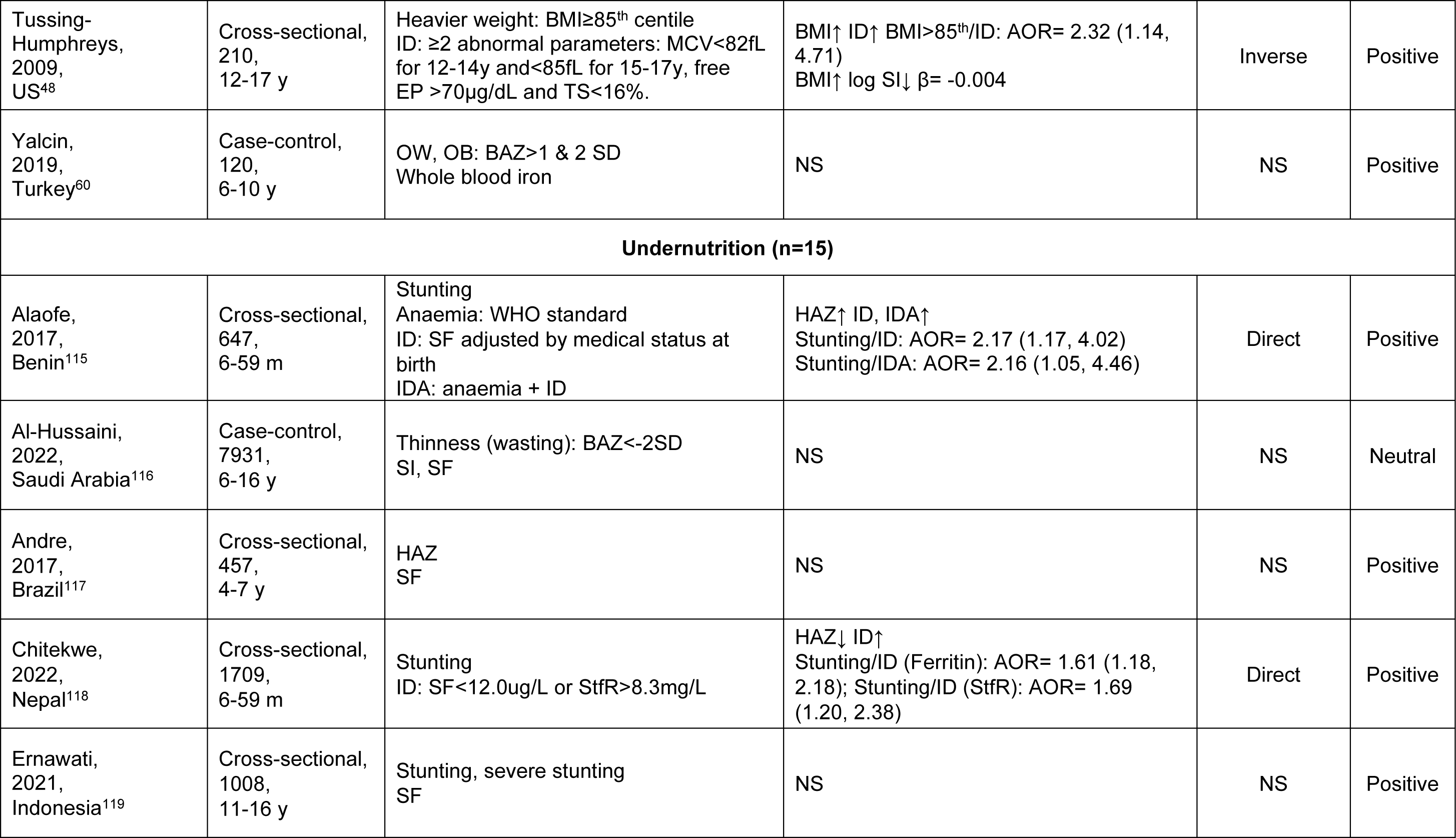
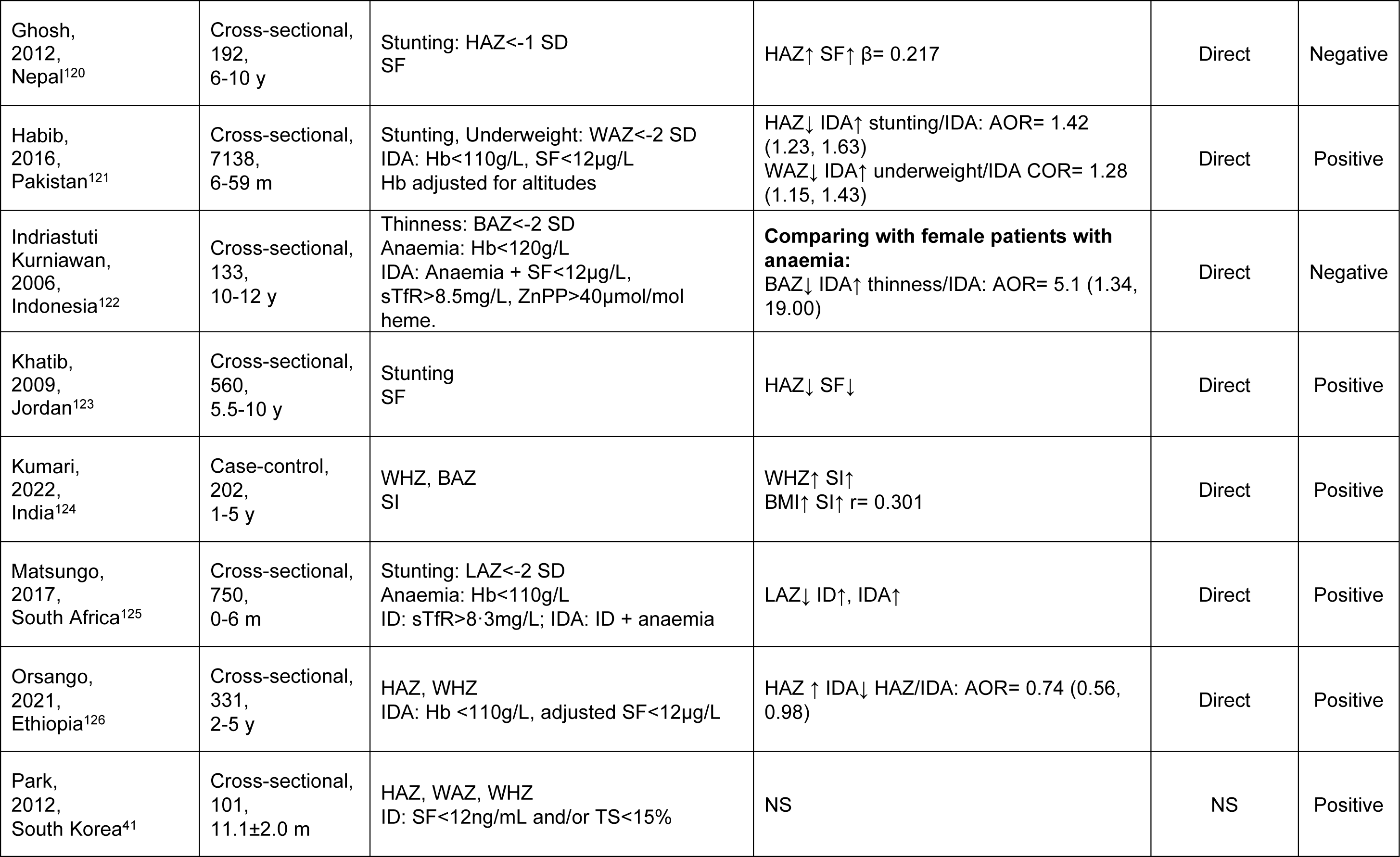
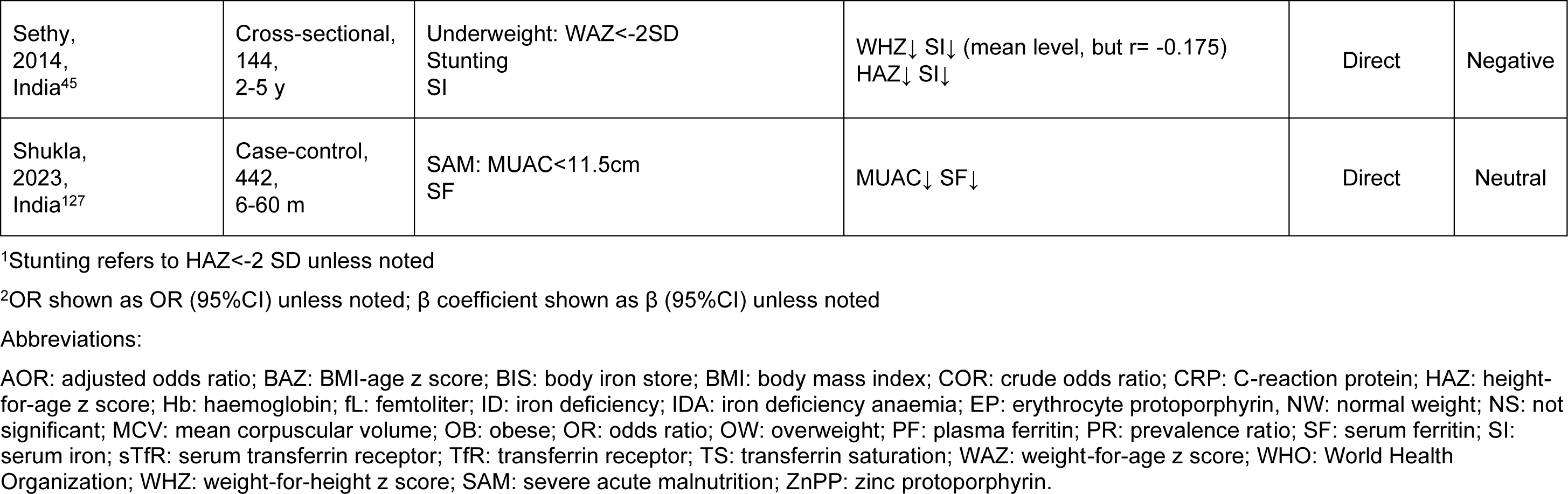
Characteristics of studies (n=46) assessing the association between iron and under- and overnutrition.

**Table 2.**
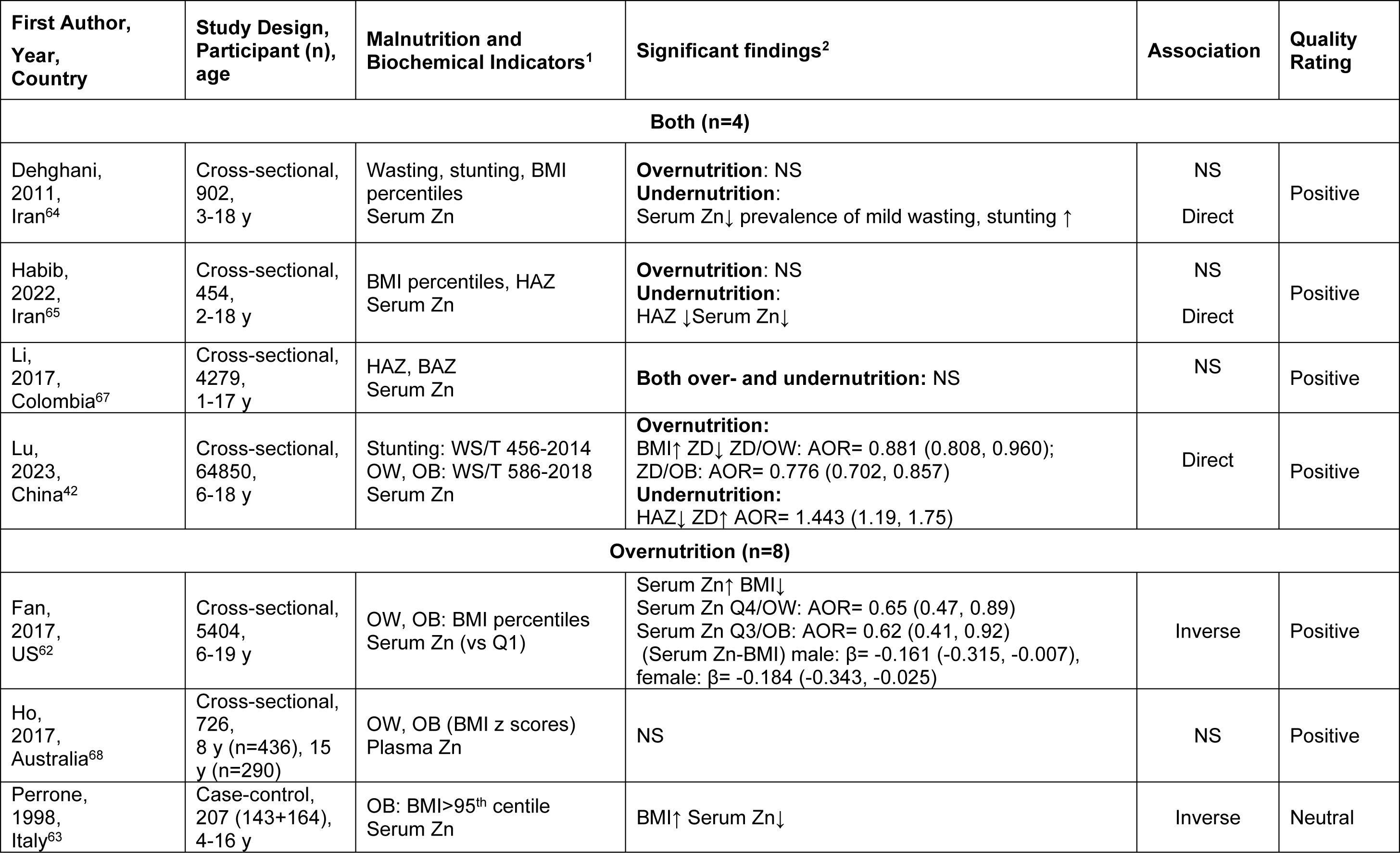
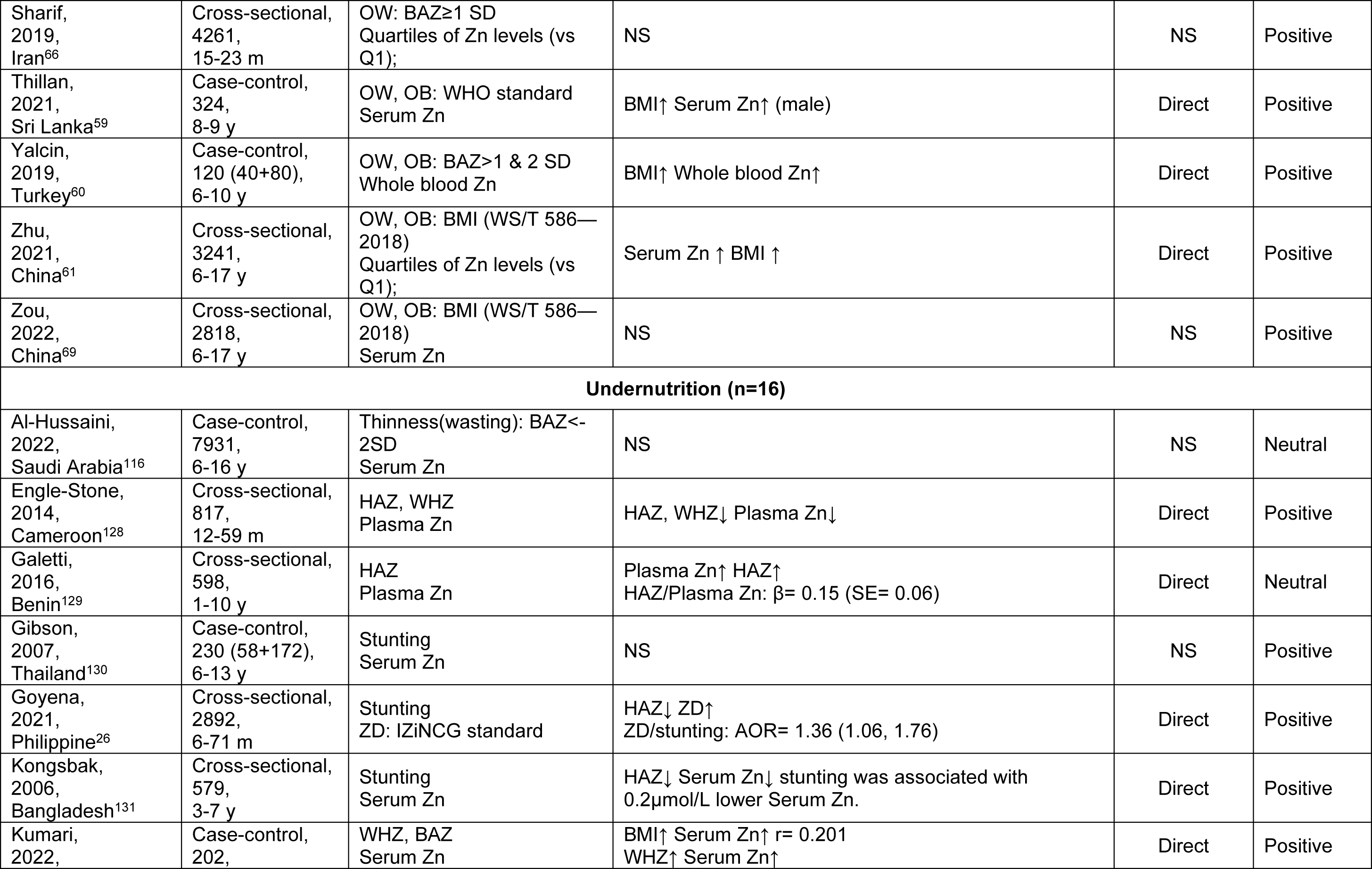
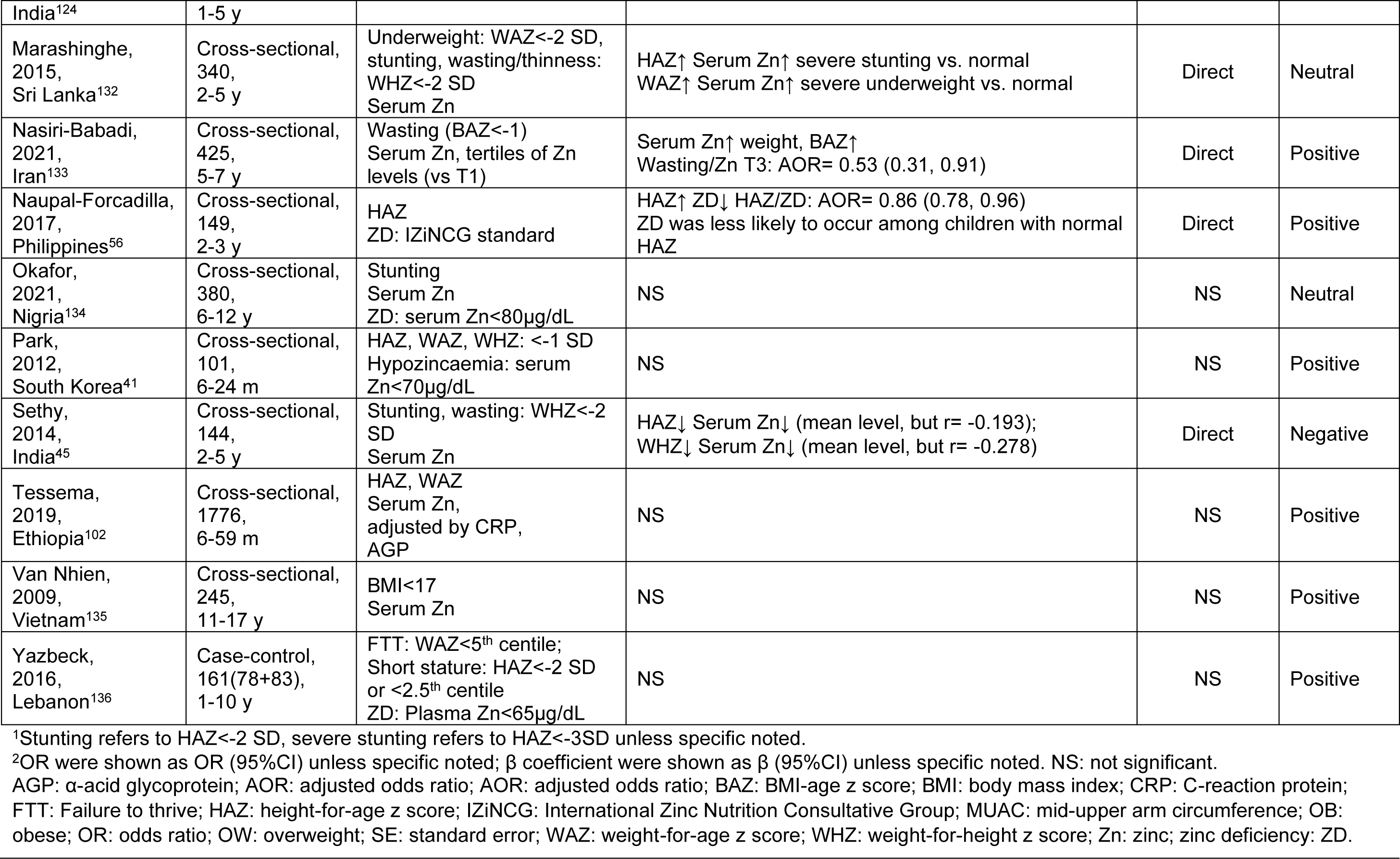
Characteristics of the studies (n=28) assessing the association between zinc and weight status.

**Table 3.** Characteristics of the studies (n=27) assessing the association between vitamin A and weight status.

| First Author, Year, Country | Study Design, Participant (n), age | Malnutrition and Biochemical Indicators<sup>1</sup> | Significant findings<sup>2</sup> | Association | Quality Rating |
| --- | --- | --- | --- | --- | --- |
| **Both (n=5)** |  |  |  |  |  |
| Dallazen, 2023, Brazil<sup>83</sup> | Cross-sectional, 1503, 12-59 m | HAZ, BAZ<br>VAD: plasma retinol<0.7µmol/L | **Overnutrition:**<br>BAZ↑ plasma retinol↓<br>**Undernutrition:**<br>HAZ↓ VAD↑ APR= 4.75 (2.10, 10.73) plasma retinol↓ | Inverse<br>Direct | Positive |
| Disalvo, 2019, Argentina<sup>78</sup> | Cross-sectional, 624, 1-6 y | WAZ, BMIz<br>Serum retinol | **Overnutrition:** BMIz↑ Serum VA↑<br>**Undernutrition:**<br>WAZ↓ Serum retinol↓ | Direct | Positive |
| Li, 2017, Colombia<sup>67</sup> | Cross-sectional, 3844, 1-17 y | HAZ, BAZ<br>Serum VA | **Both under- and overnutrition:**<br>BAZ↑ Serum VA↑ | Direct | Positive |
| Maslova, 2009, Colombia<sup>79</sup> | Cross-sectional, 2811, 5-12 y | BAZ<br>Plasma retinol | **Both under- and overnutrition:**<br>Plasma retinol↑ BAZ↑ | Direct | Positive |
| Tan, 2023, Malaysia<sup>25</sup> | Cross-sectional, 776, 7-11 y | BAZ, HAZ, WAZ<br>VAD: Both plasma retinol and RBP<0.7µmol/L | **Overnutrition:**<br>BAZ↑ retinol, RBP↑; WAZ↑ retinol, RBP↑<br>**Undernutrition:**<br>Serum VA↓ HAZ↓ VAD/stunting: COR= 2.9 (1.8, 4.7) | Direct | Positive |
| **Overnutrition (n=12)** |  |  |  |  |  |
| Cobayashi, 2013, Brazil<sup>28</sup> | Cross-sectional, 582, 0-10 y | OW: BAZ>1<br>VAD: plasma VA<20µg/dL | OW↑ VAD↑ PR= 1.97 (1.13, 3.41) for ≥5y | Inverse | Positive |
| de Souza Valente da Silva, | Cross-sectional, 471, 7-17 y | OW, OB: BMI>85<sup>th</sup> & 95<sup>th</sup> centiles<br>Low serum carotenoids:<40 µg/dL | BMI↑ serum carotenoids↓ r= -0.192<br>BMI/low serum carotenoids: β= 0.92, AOR= 2.51 (1.43, 4.39) | Inverse | Positive |
| 2007,<br>Brazil<sup>81</sup> |  |  |  |  |  |
| Gunanti,<br>2014,<br>US<sup>73</sup> | Cross-sectional,<br>1154,<br>8-15 y | OW, OB: BMI>85<sup>th</sup> & 95<sup>th</sup><br>centiles<br>Serum retinol and quartiles | BMI↑ serum retinol↑ $\beta$= 5.56 (3.36, 7.75)<br>OW/retinol quartile: AOR= 2.01(1.26, 3.22)<br>OB/retinol quartile: AOR= 2.90(1.65, 5.09) | Direct | Positive |
| Hu,<br>2001,<br>China<sup>74</sup> | Cross-sectional,<br>793,<br>0-19 y | BMI<br>Serum VA | BMI↑ VA↑ $r$= 0.26, $\beta$= 4.32±0.93<br>Weight↑ VA↑ $r$= 0.37, $\beta$= 10.39±1.85 | Direct | Positive |
| Ortega-Senovilla,<br>2019,<br>Spain<sup>75</sup> | Case-control,<br>141 (70+71),<br>6-8 y | OB (Cole et al, 2000)<br>Plasma all-trans-retinol | BMI↑ plasma retinol↑ | Direct | Neutral |
| Paes-Silva,<br>2018,<br>Brazil<sup>82</sup> | Cross-sectional,<br>411,<br>12-19 y | OW: BAZ>1;<br>Low β-carotene: Serum β-<br>carotene <0.9μmol/L | BAZ↑ β-carotene↓<br>Low β-carotene/OW, PR= 1.46 (1.2, 1.8) (male)<br>Low β-carotene/NW, PR= 1.19 (1.0, 1.4) (female) | Inverse | Positive |
| Sharif,<br>2019,<br>Iran<sup>66</sup> | Cross-sectional,<br>4261,<br>15-23 m | OW: BAZ>1<br>Quartiles of retinol levels (vs Q1) | NS | NS | Positive |
| Thillan,<br>2021,<br>Sri Lanka<sup>59</sup> | Case-control,<br>324,<br>8-9 y | OW; OB: WHO standard<br>Serum VA | NS | NS | Positive |
| Tian,<br>2022,<br>China<sup>76</sup> | Cross-sectional,<br>3025,<br>7-17 y | General OB: WS/T 586-2018<br>Central OB: WC≥90<sup>th</sup> centile<br>Quartiles of Serum VA levels (vs<br>Q1) | serum VA↑ general OB↑ Q4/general OB: AOR= 4.10<br>(2.88, 5.85)<br>serum VA↑ central OB↑ Q4/central OB: AOR= 4.14<br>(3.07, 5.56) | Direct | Positive |
| Wei,<br>2016,<br>China<sup>84</sup> | Cross-sectional,<br>1928,<br>7-11 y | OB: BMI>95<sup>th</sup> centile<br>VAD: Serum VA <20μg/dL | BMI↑ VAD/OB: AOR= 2.37 (1.59, 3.55)<br>BMI↑ Serum VA↓ | Inverse | Positive |
| Yang,<br>2015,<br>China<sup>77</sup> | Cross-sectional,<br>3457,<br>7-17 y | OW, OB: BMI>85<sup>th</sup> & 95<sup>th</sup><br>centiles<br>VAD: serum retinol<30μg/dL | BMI↑ serum retinol↑<br>VAD/NW: AOR= 1.34 (1.10, 1.63), $\beta$= 0.29 | Direct | Positive |
| Zou,<br>2022, | Cross-sectional,<br>2818 | OW; OB: WS/T 586-2018<br>VAI: Serum VA 20-30μg/dL; | serum VA↑ BMI↑<br>sufficient VA/OW: AOR= 1.55 (1.19, 2.02) | Direct | Positive |
| China<sup>69</sup> | 6-17 y | VAD: Serum VA<20µg/dL |  |  |  |
| **Undernutrition (n=10)** |  |  |  |  |  |
| Adamu, 2016, Nigeria<sup>137</sup> | Case-control, 550 (275+275), 6 m-5 y | WHZ<br>Serum retinol | WHZ↓ Serum VA↓ | Direct | Neutral |
| Ahmed, 2006, Bangladesh<sup>138</sup> | Cross-sectional, 381, 11-16 y | BMI<br>Serum VA | BMI↓ Serum VA↓ r= 0.33, β= 0.26 | Direct | Positive |
| Alaofe, 2017, Benin<sup>115</sup> | Cross-sectional, 647 children, 6-59 m | Stunting<br>VAD: serum retinol<20µg/dL, corrected by infection | NS | NS | Positive |
| Ernawati, 2021, Indonesia<sup>119</sup> | Cross-sectional, 1008, 11-16 y | Stunting; severe stunting<br>Serum retinol | HAZ↓ serum retinol↓ | Direct | Positive |
| Khatib, 2009, Jordan<sup>123</sup> | Cross-sectional, 560, 5.5-10 y | BMI; stunting<br>Serum VA | BMI↓ serum VA↓ r= 0.114 | Direct | Positive |
| Kurugol, 2000, Turkey<sup>139</sup> | Cross-sectional, 160, 6-59 m | Stunting<br>Serum retinol | HAZ↓ Serum retinol↓ | Direct | Neutral |
| Marashinghe, 2015, Sri Lanka<sup>132</sup> | Cross-sectional, 340, 2-5 y | Underweight: WAZ<-2; stunting; wasting/thinness: WHZ<-2<br>Serum VA | NS | NS | Neutral |
| Oso, 2003, Nigeria<sup>140</sup> | Cross-sectional, 213, 6 m-6 y | Stunting<br>Serum retinol | HAZ↓ serum retinol↓ | Direct | Negative |
| Samba, 2006, Congo<sup>141</sup> | Cross-sectional, 5722, 0-71 m | WHZ <-2 SD<br>Serum retinol | WHZ↓ Serum retinol↓ | Direct | Positive |
| Ssentongo, 2020, Uganda<sup>27</sup> | Cross-sectional, 4765, 6–59 m | Stunting; severe stunting<br>VAD: RBP<0.83µmol/L, adjusted by CRP | RBP↓ HAZ↓<br>VAD/stunting: AOR= 1.43 (1.08, 1.89)<br>VAD/severe stunting: AOR= 1.64 (1.14, 2.35) | Direct | Positive |

<sup>1</sup>Stunting refers to HAZ<-2 SD, severe stunting refers to HAZ<-3SD unless specific noted.
<sup>2</sup>OR were shown as OR (95%CI) unless specific noted; β coefficient were shown as β (95%CI) unless specific noted.
AOR: adjusted odds ratio; BAZ: BMI-age z score; BMI: body mass index; CRP: C-reaction protein; HAZ: height-for-age z score; NS: not significant; NW: normal weight; OB: obese; OR: odds ratio; OW: overweight; RBP: retinol binding protein; VAD: vitamin A deficiency; WAZ: weight-for-age z score; WHZ: weight-for-height z score.

### Associations between iron and weight status

The characteristics and findings of the studies that reported associations between iron status and weight status in children and young people (n=46) are summarised in **Table 1**. Ten studies assessed both sides of malnutrition, whereas 21 studies focused on overnutrition, and 15 studies focused on undernutrition. Although a broad range of biomarkers for ID and overnutrition were used by the different studies with many using more than one, and most at least utilised serum ferritin as a minimum to assess iron status (**Table 1**). In 2020 the WHO revised their guidance on the use of ferritin to assess iron status in individuals and populations, providing thresholds of <15μg/L for healthy individuals above 5 years of age and <70μg/L for individuals with infection or inflammation (<12μg/L or <30μg/L are used for children under 5). While current WHO guidance is that ferritin, both an iron storage protein and an acute phase reactant associated with inflammation^43^, should not be used alone to diagnose ID without other iron profile biomarkers or corrections for inflammation^44^, a minority of studies did rely on ferritin alone^25 49 100–102^. Likewise, serum iron alone should not be used in isolation to diagnose ID as serum iron levels will be reduced in the context of infection or inflammation^98^, but was used in a handful of studies^43 46 47^. In risk of bias assessment, most studies were found to be of positive quality (38 out of 47; **Table 1, Table S5**). Only one study was found of negative quality^45^, with no appropriate statistical method and adequate adjustments the most common reason for studies being rated either neutral or negative quality (**Table S5**).

A total of 16 records reported OR from 7 eligible studies enabling our calculation of the pooled OR for ID with overnutrition (overweight or obesity) using the normal weight groups as reference. These studies assessed overnutrition and ID in North American (US^46–48^ and Mexico^49^) and European (Spain^50^ and Greece^51 52^) populations; and, in risk of bias assessment, all were assessed as positive (good) quality. The random-effects model meta-analysis indicated a pooled OR (95% CI) of 1.51 (1.20, 1.82), p<0.0001 for ID with overnutrition (**Figure 3**). Heterogeneity was moderate with a I^2^ value of 40.73%, likely a result of the diversity of ethnicities, ages and genders studied; as well as the broad range of biomarkers for ID and overnutrition used by the different studies. Subgroup analyses suggested increased risk of ID in those living with obesity OR (95%CI): 1.88 (1.33, 2.43), p<0.0001 compared to overweight OR (95%CI): 1.31(0.98, 1.64), p<0.0001; however between group differences were not statistically different (p=0.08), likely driven by greater heterogeneity in the overweight versus obesity data (I^2^ 40.46% versus 20.0%, **Figure 3**).

**Figure 3.**
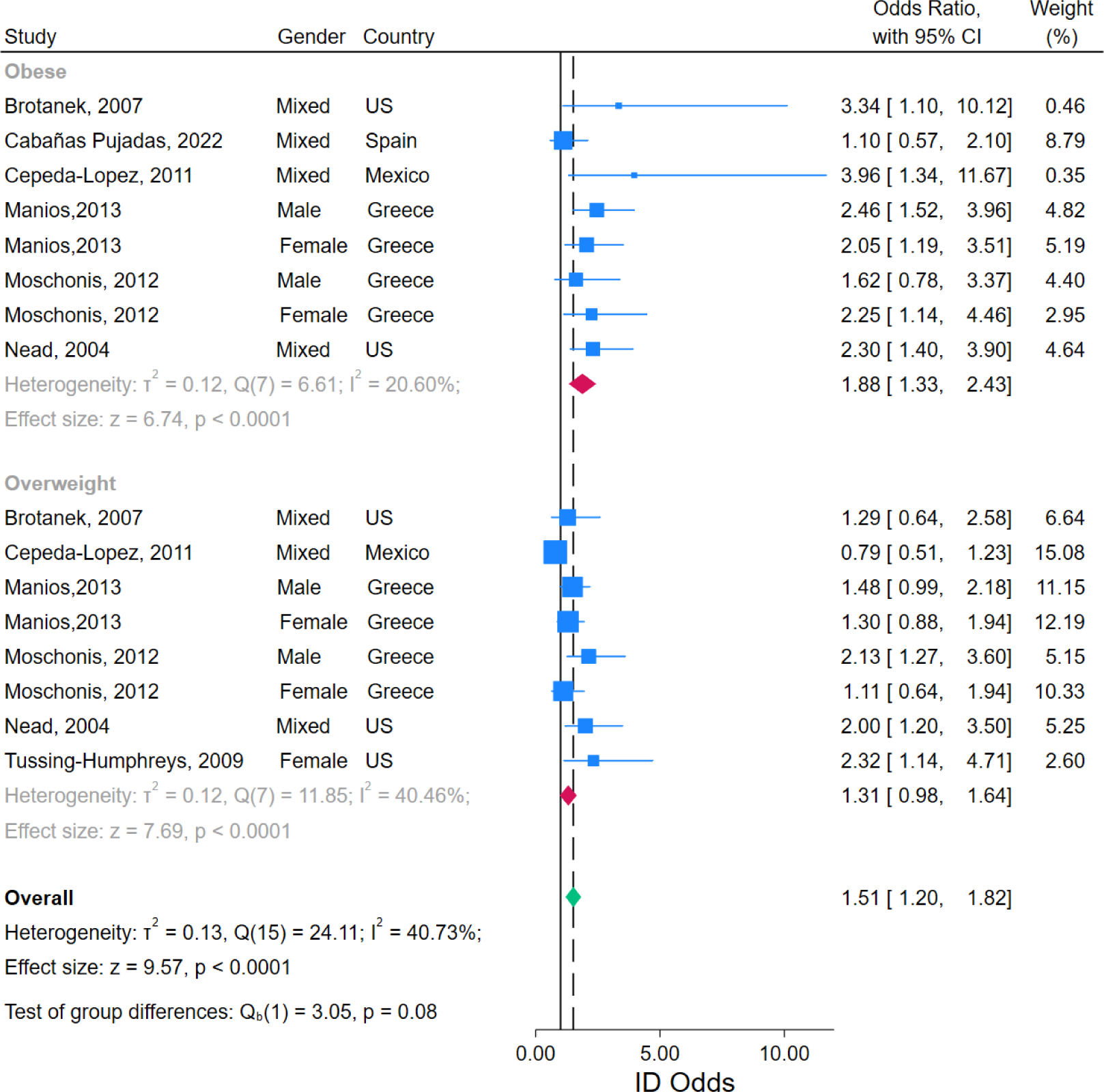
Forest plot of reported associations between overnutrition and odds of iron deficiency. REML: Random effects restricted maximum likelihood; CI: confidence interval.

Similarly, no differences were observed between studies that reported data separately for males and females versus those that reported data from mixed-gender populations (p=0.47, **Figure S1)**. Funnel plot analyses suggested some asymmetry driven by two outliers (**Figure S2)**. These studies reported higher prevalence of ID in both overweight and obese mixed-gender populations, with much larger standard errors observed in the (fewer) children with obesity (**Figure 3**)^46 49^. As they contributed only small weights to the meta-analysis (Brotanek et al: 0.46%, Cepeda-Lopez et al: 0.35%), this is a small-study effect rather than non-reporting bias, and indeed funnel analyses with fewer than 10 studies should be interpreted cautiously^53^. Sensitivity analyses indicated robustness in the overall results as the effect size was not significantly changed after removing any of the studies (**Figure S3**).

Given that the meta-analysis incorporated only a small subset (7 of 46) of iron studies, to synthesize the findings across all studies, we examined the numbers of participants in the reports concluding either direct, inverse, or no significant associations between iron deficiency and nutrition status (**Figure 4A**). These data illustrate surprising heterogeneity in the conclusions of individual studies and show where individual studies contributed disproportionally larger populations. In the case of iron and undernutrition, while 13 reports that included 15,335 participants found a direct association (i.e., improved iron status as move from undernutrition to normal bodyweight), a surprising 10 studies that included 30,705 participants found no association, and 2 concluded an inverse association (**Figure 4A**). Notably, the 2 studies that found an inverse association had reported linear regression between iron and both under- and overnutrition (BMI 14-32^54^ and BMI-z score between −4 and 2^55^). However, the associated scatter plots suggest piecewise regression would be more appropriate as very low ferritin values were observed in both the lowest and highest quintiles of nutrition status. While the data for overnutrition appear clearer, with 19 reports including 27,894 participants concluding an inverse relationship (i.e., worsening iron status as weight status moves from normal to overweight and obesity), nonetheless 9 reports with 8,884 participants concluded a direct association and 3 found no association (**Figure 4A**).

**Figure 4.**
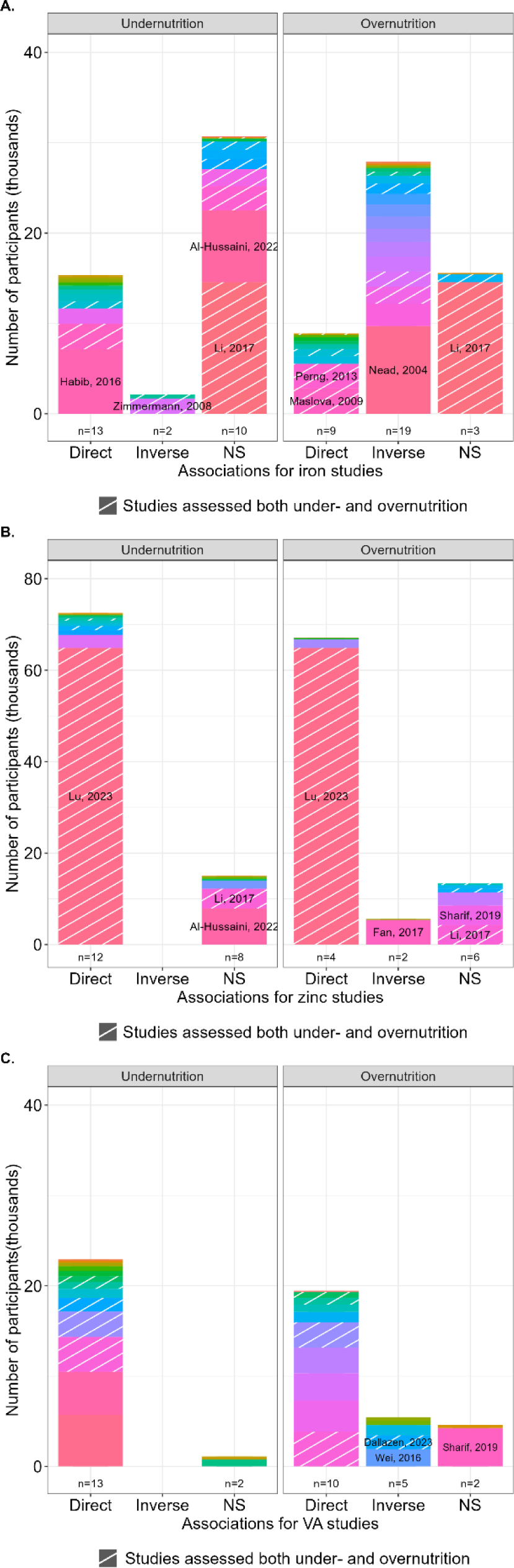
Summary stacked bar plots showing participant numbers, grouped by the direction of associations, in studies examining overnutrition or undernutrition**. A**. Iron, **B.** Zinc, and **C.** Vitamin A. Studies where participants represented >25% of the total population in each category were labelled.

While these conflicting results likely reflect different populations studied and the challenges of ID diagnosis, nonetheless, we conclude the data collectively suggest an inverted U-shaped relationship between iron status and bodyweight; with children and young people living with overnutrition having a higher risk [pooled OR (95% CI): 1.51 (1.20, 1.82)] of iron deficiency alongside those with undernutrition.

### Associations between zinc and weight status

The 28 studies that examined zinc status and either under- or overnutrition were similarly diverse in their approaches and findings (**Table 2**). While all 28 studies measured either plasma or serum zinc, how they were compared statistically (e.g. mean, median, quartile (Q) Q2-4 versus Q1), varied between studies, and precluded any meta-analysis. We note only 2 studies^26 56^ used international consensus-based thresholds (57-74 μg/dL depending on age group, sex, time of day and time since last meal^57^) for defining deficiency, which may explain some of the heterogeneity in conclusions between studies here. In addition, although plasma or serum zinc concentration is the most used biomarker of zinc status, there are well described issues with sensitivity and specificity, as well as challenges with sample collection and storage, that render the assessment of dietary zinc status in humans challenging^58^. In risk of bias assessment, most studies (22 out of 28) received a positive quality rating.

Although a direct relationship between zinc status and undernutrition was concluded by the majority of studies (12 of 20 accounting for 72,537 participants; **Figure 4B**), the results from reports on overnutrition were more contradictory. While 4 studies concluded direct associations between zinc levels and BMI^42 59–61^, 2 found inverse relationships^62 63^ and 6 studies, accounting for 13,440 participants in 4 countries (3 from Iran^64–66^, and 1 from Colombia^67^, Australia^68^, and China ^69^), concluded no association between zinc status and overweight (**Figure 4B**). Among the studies reporting direct associations with both under- and overnutrition was an exceptionally large population study from China with >64,000 participants^42^. While in this cohort stunting conferred a higher risk of zinc deficiency [OR (95%CI): 1.44 (1.19-1.75)], and overweight and obesity appeared to confer protection [overweight: 0.88 (0.84, 0.96); obesity: 0.78 (0.70, 0.86)], serum zinc values were not hugely different between groups and within adequate population ranges^57 70^; with 86.0μg/dL (77.0, 96.0) reported for children with stunting, and 91.0μg/dL (82.0, 101.0) for children living with obesity^42^. Relatedly, in the larger (n=5,404 American children aged 5-19) the overall population mean and (range) of zinc levels were similarly adequate, 82.7μg/dL (38.9, 198.6)^62^.

In sum, while the data were more limited for zinc, they suggest an exponential plateau curve relationship between zinc status and bodyweight, with children and young people living with severe undernutrition having the highest risk for zinc deficiencies, and no evidence for a higher risk for zinc deficiency with overnutrition.

### Associations between vitamin A and weight status

The studies that investigated vitamin A status in relation to body weight were similarly heterogenous in their approaches to assessing both vitamin A and nutrition status (**Table 3**). Vitamin A was commonly assessed by measuring plasma/serum retinol levels, although some studies measured either serum carotenoids, β-carotene, or retinal binding protein (RBP) levels. However, plasma/serum retinol concentration is under tight homeostatic control and does not reflect vitamin A status until body stores are extremely low or very high^71^. Where low serum retinol was defined, WHO cut-offs (retinol<0.7μmol/L, equivalent to 20μg/dL^72^) were most often but not always used. These values are not sensitive to moderate vitamin A deficiencies, and like with zinc, studies often examined results by comparing bottom and top quartiles of the biomarkers measured. As with the zinc studies, variability in the outcomes measured and statistical approaches between studies prevented us from performing meta-analysis, and the majority of studies received positive quality ratings (23 out of 27; **Table 3, Table S5**).

While a direct relationship between undernutrition and vitamin A deficiency was clear with 13 of 15 studies concluding a direct association, other 2 reported no association (**Figure 4C**), the data for associations between overnutrition and vitamin A deficiency were more contradictory. Although in favour of a direct relationship between vitamin A status (n=10 of 17 studies, accounting for 19,443 participants), 5 found an inverse association and 2 found no association (**Figure 4C**). In the 10 studies that found a direct relationship, these were reported in a variety of ways, such as: lower risk of VAD in the context of overweight or obese^69 73–77^, or elevated serum levels of vitamin A or plasma retinol in children with overweight or obesity^67 78–80^. Interestingly, of the 5 studies that concluded an inverse association between overnutrition and vitamin A deficiency, 4 focused on Brazilian children. The associations were reported either as increased risk of VAD^28^, or risk of low carotenoids^81 82^, for overweight children aged 5-19 with around 500 participants in each study. While in one case, low plasma retinol (<0·70 μmol/L) was associated with overweight in Brazilian children aged 12–59 months old (n=1503)^83^. The fifth, non-Brazilian, study assessed 7–11 years old children (n=1928) in Chongqing, China^84^, and found that obesity increased the risk of VAD, OR (95%CI):2.37 (1.59, 3.55).

In the context of the tight homeostatic mechanisms for plasma retinol levels^71^ we conclude that similar to zinc, the data suggest an exponential plateau curve relationship between vitamin A status and bodyweight, with children and young people living with severe undernutrition having the highest risk for vitamin A deficiencies, and no evidence for a higher risk for zinc deficiency with overnutrition.

Lastly, regional differences were observed both in terms of which micronutrients were more frequently investigated and whether populations with under- or overnutrition or both were investigated (**Figure 5**). Studies from North America and Europe (**Figure 5A**) focused entirely on overnutrition and largely on iron. Whereas reports from the Western Pacific (**Figure 5B**), Asia (**Figure 5C**) and Latin America (**Figure 5D**) assessed both under- and overnutrition, most studies in Africa (**Figure 5E**) focused on undernutrition.

**Figure 5.**
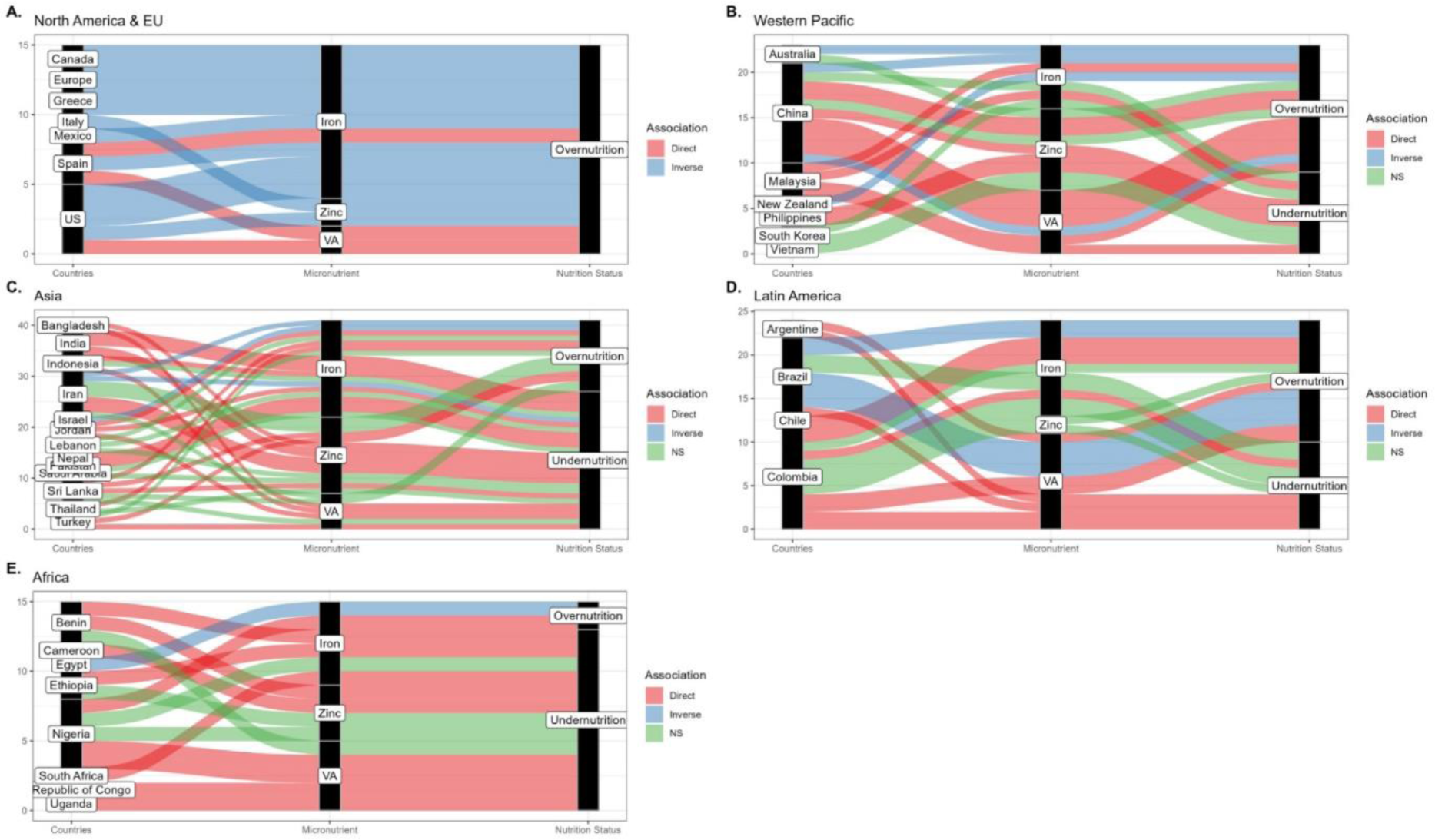
Sankey plots illustrating regional differences in study characteristics and conclusions. **A.** North America and Europe, **B.** Western Pacific, **C.** Asia (excluding Western Pacific), **E.** Latin America, **F.** Africa. Each line represents a reported association for different micronutrient and nutritional status in different country. The size of each sub-columns represented the proportion of each category.

## DISCUSSION

To our knowledge, this is the first systematic review of the associations between weight status in children and young people and iron, zinc, and vitamin A deficiencies, to concurrently examine associations with both under- and overweight, i.e., the double burden of malnutrition. Notably, our results suggest an inverted U-shaped relationship between iron status and bodyweight, with both under- and overnutrition increasing risk for ID. In meta-analyses children and young people living with overnutrition had a higher risk [pooled OR (95% CI): 1.51 (1.20, 1.82)] of iron deficiency, with those living with obesity having even higher risk [pooled OR (95% CI): 1.88 (1.33, 2.43)] compared to those with normal weight. In contrast, zinc and vitamin A deficiencies were most observed in children and young people with undernutrition. While in aggregate, the studies underscore the increased risks for iron, zinc, and vitamin A deficiencies with undernutrition in childhood and adolescence, there was more heterogeneity in study conclusions (i.e., several studies finding no relationship) for iron and zinc. Such heterogeneity in part relates to the methodological challenges of assessing the status of these micronutrients in humans. Indeed, our results highlight how variable the approaches to measuring and reporting dietary nutrient deficiencies have been, as well as regional data gaps, with important implications for future study design.

Excess adiposity is associated with low-grade inflammation, widely proposed to be the mechanism behind reduced iron status observed in adults living with obesity^85 86^. With the expansion of adipose tissue comes macrophage infiltration and release of inflammatory cytokines such as IL-6, that stimulate the synthesis of acute phase proteins such as ferritin and hepcidin, the main regulator of systemic iron homoeostasis^85^. Hepcidin prevents iron absorption from the enterocytes and iron release from splenic macrophages, thereby reducing circulating iron levels overall^86^. In acute infection, this serves to deprive a pathogen of iron^87^, but in the context of chronic disease with prolonged immune activation can lead to anaemia of chronic disease^88^. The results of our meta-analysis, showing a higher risk of iron deficiency with overnutrition, particularly in children and young people living with obesity, are consistent with those seen in adults^16^; and suggest inflammation as a factor, however inflammatory markers were only rarely measured.

Inflammatory status is particularly relevant as ferritin was the primary blood biomarker of iron status used, consistently found higher in obese groups compared to children with normal weight and direct linear association reported in multiple studies^25 50 55 59 79 89–99^. However, as ferritin is an acute phase reactant, inflammation can obscure ID^100^, therefore when the WHO reviewed their guidance on the use of ferritin to assess iron status in 2020^44^, they made the strong recommendation that in areas of widespread infection or inflammation, serum ferritin should be measured alongside both C-reactive protein (CRP) and α-1 acid glycoprotein (AGP), which reflect different phases of the acute phase response from acute infection to chronic inflammation. In addition, they recommend that thresholds to define ID in individuals with infection or inflammation should be raised to 30 μg/L for children under five and 70 μg/L serum ferritin for all age groups over five^44^. However, very few studies (n=4 out of 23 studies that used ferritin with or without other biomarkers) to define ID) measured and excluded participants with elevated CRP level^94 98 99 101^. Only one study measured AGP^102^ and only 2 studies from Colombia^79^ and Brazil^103^ utilised the higher thresholds for ferritin to define ID in the context of inflammation. Interestingly, the WHO serum ferritin concentration thresholds were based on expert opinion^44^, and a recent analysis of serial cross-sectional NHANES data concluded thresholds for ferritin for iron-deficient erythropoiesis should be higher, at least for healthy American children and women^104^. Specifically, from regression models of the distributions of haemoglobin and sTfR with serum ferritin in 2569 and 7498 healthy children under five and women (15-49 years old) they derived 20 μg/L for children under five and 25 μg/L in women as physiologically based serum ferritin thresholds for ID. These data suggest that the global burden of iron deficiency in children and young people may well be underestimated.

In contrast to iron, our results show that zinc and vitamin A deficiencies were most observed in undernutrition, with limited evidence for a higher risk for deficiency with overnutrition. While variability in the outcomes measured and statistical approaches between studies precluded meta-analyses, critical review of the data in aggregate suggest an exponential plateau curve relationships between zinc or vitamin A status and bodyweight, with children and young people living with severe undernutrition having the highest risk for these micronutrient deficiencies. Nonetheless, considerable uncertainty in the evidence base remains, particularly for overnutrition. This is not least of all because of efficient homeostatic mechanisms that buffer plasma zinc and vitamin A levels to either dietary deficiency or excess, thereby maintaining plasma concentrations within a narrow range, and preventing toxicity in the case of vitamin A^71 105^. Therefore, more moderate deficiencies in overnutrition may be masked. In addition, mechanistically there are complicated interactions between the micronutrients themselves (e.g. vitamin A deficiency impairs iron mobilisation and vitamin A supplementation will improve haemoglobin concentrations^105^), and between overnutrition, inflammation and micronutrient metabolism.

Indeed, our comprehensive review highlights the challenges of micronutrient assessment at an individual and population level with important implications for future study design and reporting in the context of the double burden of malnutrition. The most commonly used biomarkers of micronutrient status (plasma/serum ferritin, zinc, and retinol levels) are sensitive to a variety of stimuli, including infection and inflammation^72^, and a primary recommendation for future work investigating MNDs in either under- or overnutrition is that CRP and AGP are routinely measured, and inflammation accounted for in line with current expert guidelines. While in the case of iron, ferritin increases with inflammation and ID may be underestimated and thresholds for deficiency should be raised^106^, in the cases of zinc and vitamin A, deficiencies may be overestimated as both plasma/serum zinc and retinol are, at least transiently, lowered in acute infection or inflammation^107 108^. Conversely, additional challenges with zinc measurement exist as careful sample collection and handling is critical to prevent haemolysis or contamination from adventitious zinc in the environment^57^, which may mask deficiencies, a potential confounder in the studies that surprisingly did not find zinc deficiency associated with undernutrition. If CRP and AGP are measured, inflammation can be adjusted for by either exclusion, the use of correction factors, or linear regression approaches recently proposed by the Biomarkers Reflecting Inflammation and Nutritional Determinants of Anaemia (BRINDA) project^109^. Regression approaches have now been systematically developed and used to adjust for ferritin^110^, zinc^108^ and vitamin A^107^ values on a continuous scale.

In addition, our work highlights that many studies did not apply consensus-based cut-offs for defining either iron, zinc or vitamin A deficiencies but rather compared lower to upper quartiles of plasma/serum biomarkers within their populations, often not specifying the measured ranges of biomarkers within quartiles. Such calculations may overestimate risks of deficiencies in largely replete populations, and their widespread use may explain some of the seeming contradictory conclusions. Lastly, our data highlights regional data gaps and differences in study focus, with most studies from Africa and Asia focused on undernutrition and those from North America and Europe focused entirely on overnutrition. This is concerning both Africa and Asia experienced dramatic increases in the number of overweight children under 5 between 2000 and 2017 (from 6.6 to 9.7 million children in Africa and from 13.9 to 17.5 million children in Asia)^111^. Moreover, the regions of Africa and Asia have the highest double burden of malnutrition with large numbers of stunted and wasted children, and the number of stunted children under 5 having increased from 50.6 to 58.7 million children in Africa^111^. These stark data underscore that the investigation of MNDs in relation to the double burden of malnutrition remains critically important for child health.

This is the first study to concurrently, comprehensively review the associations between iron, zinc, and vitamin A deficiencies and the double burden of malnutrition in children and young people. In general, undernutrition groups were reported to have lower plasma/serum ferritin, zinc, and retinol levels, or found to have higher risks for iron, zinc, and vitamin A deficiencies. Evidence gaps notwithstanding, we conclude that overnutrition was associated with iron deficiency, but not zinc or vitamin A deficiencies. Strengths of this work included our comprehensive search strategy with robust inclusion and exclusion criteria and *a priori* registration of detailed review protocol in line with PRISMA guidelines^32^ and utilising the PICO approach^112^. In addition, this systematic review included populations across the globe (including North American and Europe, Latin American, Western Pacific, Asia and Africa continents), which allowed us to identify regional data gaps and differences in term of their nutritional outcomes or micronutrients of interests and the associations reported. While we excluded studies with small sample size (<100) and those that employed convenience sampling methods to reduce the bias; this review has also highlighted the importance of addressing the challenges and weaknesses in study design, analytical methods, and diagnostic criteria for micronutrient deficiencies, to ensure the validity and reliability of data in future study for interpretation. Nonetheless, this work also has some limitations. Firstly, all observational studies are limited by confounding and causality should not be inferred. Second, only 4 databases (Medline, Scopus, Embase and Cochrane) were searched, and only studies written in English were included, which may have limited the scope. Last and not least of all, MND status assessment is not trivial in humans, and the underlying studies included in this review were heterogeneous in the populations studied and approaches taken, conferring some limitations in data interpretation.

## SUMMARY

Iron, zinc and vitamin A deficiencies were commonly associated with undernutrition in children and young people. Overnutrition increased the risk of iron deficiency, but not zinc or vitamin A deficiencies, with an inverted U-shaped relationship observed between iron status and bodyweight. Heterogeneity between studies was attributable to the diversity of participants, diagnostic criteria, and approaches to data analysis. Inflammation status was rarely adequately assessed, and we conclude the burden of iron deficiency may well be under-recognised, particularly in children and young people living with overnutrition.

### Author Contributions

Conceptualization, X.T., P.Y.T., Y.Y.G., and J.B.M.; Data curation, formal analysis, investigation, and methodology, X.T. and P.Y.T.; Formal analysis, X.T. and P.Y.T.; Supervision, Y.Y.G., and J.B.M.; Validation, X.T. and P.Y.T.; Writing—original draft, X.T.; Writing—review and editing, X.T., P.Y.T., Y.Y.G., and J.B.M. All authors approved the final manuscript.

### Funding

This research was funded by the project titled “Addressing micronutrient deficiencies associated with the double burden of childhood malnutrition in China, a combined food system framework; BB/T008989/1” from UK Biotechnology and Biological Sciences Research Council (BBSRC).

## Data Availability

All data produced in the present work are contained in the manuscript

