## Supplemental Information for "Overnutrition is a risk factor for iron deficiency in children and young people: a systematic review and meta-analysis of micronutrient deficiencies and the double burden of malnutrition"

**Table S1  Search strategy for Medline.**

| # ▲ | Searches | Results |
|---|---|---|
| 1 | Anaemia, Iron-Deficiency/bl, di, ep, et, me [Blood, Diagnosis, Epidemiology, Etiology, Metabolism] | 7101 |
| 2 | Iron/bl, df [Blood, Deficiency] | 17580 |
| 3 | ((iron or ferritin or hemoglobin) and (serum or blood or plasma) and (inadequa* or insufficien* or deficien*)).ti,ab,kw. | 16221 |
| 4 | 1 or 2 or 3 | 33690 |
| 5 | Zinc/bl, df [Blood, Deficiency] | 11105 |
| 6 | (zinc and (serum or blood or plasma) and (inadequa* or insufficien* or deficien*)).ti,ab,kw. | 4505 |
| 7 | 5 or 6 | 13387 |
| 8 | Vitamin A Deficiency/bl, di, ep, et, pc [Blood, Diagnosis, Epidemiology, Etiology, Prevention & Control] | 2240 |
| 9 | Vitamin A/bl [Blood] | 5182 |
| 10 | ((retinol or "vitamin A") and (serum or blood or plasma) and (inadequa* or insufficien* or deficien*)).ti,ab,kw. | 2923 |
| 11 | 8 or 9 or 10 | 8055 |
| 12 | 4 or 7 or 11 | 51425 |
| 13 | Obesity/bl, di, ep, et, me, pc [Blood, Diagnosis, Epidemiology, Etiology, Metabolism, Prevention & Control] | 98565 |
| 14 | Obesity, Abdominal/bl, di, ep, me, pc [Blood, Diagnosis, Epidemiology, Metabolism, Prevention & Control] | 2530 |
| 15 | Pediatric Obesity/bl, di, ep, et, me, pc [Blood, Diagnosis, Epidemiology, Etiology, Metabolism, Prevention & Control] | 6908 |
| 16 | Overweight/bl, di, ep, et, me, pc [Blood, Diagnosis, Epidemiology, Etiology, Metabolism, Prevention & Control] | 13827 |
| 17 | Malnutrition/bl, di, ep, et, me, pc [Blood, Diagnosis, Epidemiology, Etiology, Metabolism, Prevention & Control] | 9368 |
| 18 | Protein-Energy Malnutrition/bl, di, ep, et, me, pc [Blood, Diagnosis, Epidemiology, Etiology, Metabolism, Prevention & Control] | 4276 |
| 19 | Thinness/bl, di, ep, et, pc [Blood, Diagnosis, Epidemiology, Etiology, Prevention & Control] | 2585 |
| 20 | Body Mass Index/ | 130582 |
| 21 | Anthropometry/ | 39336 |
| 22 | (malnutrition or malnourish* or overnutrition or undernutrition or obese or obesity or overweight or adiposity or stunting or stunted or underweight or (wasting adj2 disease) or wasted or "body mass index" or BMI or anthropometr*).ti,ab,kw. | 586872 |
| 23 | 13 or 14 or 15 or 16 or 17 or 18 or 19 or 20 or 21 or 22 | 644487 |
| 24 | 12 and 23 | 4893 |
| 25 | limit 24 to english | 4565 |
| 26 | exp adolescent/ or exp child/ or exp infant/ or (infant disease* or childhood disease*).ti,ab,kf. or (adolescen* or babies or baby or boy? or boyfriend or boyhood or girlfriend or girlhood or child* or girl? or infan* or juvenil* or kid? or minors or minors* or neonat* or neo-nat* or newborn* or new-born* or paediatric* or peadiatric* or pediatric* or perinat* or preschool* or puber* or pubescen* or school* or teen* or toddler? or underage? or under-age? or youth*).ti,ab,kf. or (pediatric* or | 5094920 |

| | | |
|---|---|---|
| | paediatric* or infan* or child* or adolescen* or young).jn,jw. or (pediatric* or paediatric* or infan* or child* or adolescen* or young).in. | |
| 27 | 25 and 26 | 2549 |
| 28 | limit 27 to (address or case reports or comment or editorial or letter or meta analysis or news or "review" or "systematic review") | 302 |
| 29 | 27 not 28 | 2247 |

**Table S2 Search strategy for Embase.**

| # ▲ | Searches | Results |
|---|---|---|
| 1 | iron/ec [Endogenous Compound] | 20090 |
| 2 | iron deficiency/co, di, dm, ep, et, pc [Complication, Diagnosis, Disease Management, Epidemiology, Etiology, Prevention] | 2292 |
| 3 | iron deficiency anaemia/co, di, dm, ep, et, pc [Complication, Diagnosis, Disease Management, Epidemiology, Etiology, Prevention] | 6259 |
| 4 | ((iron or ferritin or hemoglobin) and (serum or blood or plasma) and (inadequa* or insufficien* or deficien*)).ti,ab,kw. | 24033 |
| 5 | or/1-4 | 46391 |
| 6 | zinc/ec [Endogenous Compound] | 8578 |
| 7 | zinc deficiency/co, di, ep, et, pc [Complication, Diagnosis, Epidemiology, Etiology, Prevention] | 825 |
| 8 | (zinc and (serum or blood or plasma) and (inadequa* or insufficien* or deficien*)).ti,ab,kw. | 5061 |
| 9 | or/6-8 | 12886 |
| 10 | retinol/ec [Endogenous Compound] | 4437 |
| 11 | retinol deficiency/co, di, ep, et, pc [Complication, Diagnosis, Epidemiology, Etiology, Prevention] | 1424 |
| 12 | ((retinol or "vitamin A") and (serum or blood or plasma) and (inadequa* or insufficien* or deficien*)).ti,ab,kw. | 2905 |
| 13 | or/10-12 | 7461 |
| 14 | 5 or 9 or 13 | 60740 |
| 15 | overnutrition/di, dm, ep, et, pc [Diagnosis, Disease Management, Epidemiology, Etiology, Prevention] | 234 |
| 16 | obesity/di, dm, ep, et, pc [Diagnosis, Disease Management, Epidemiology, Etiology, Prevention] | 47924 |
| 17 | abdominal obesity/di, dm, ep, et, pc [Diagnosis, Disease Management, Epidemiology, Etiology, Prevention] | 1226 |
| 18 | childhood obesity/di, dm, ep, et, pc [Diagnosis, Disease Management, Epidemiology, Etiology, Prevention] | 3482 |
| 19 | malnutrition/di, dm, ep, et, pc [Diagnosis, Disease Management, Epidemiology, Etiology, Prevention] | 7850 |
| 20 | protein calorie malnutrition/di, dm, ep, et, pc [Diagnosis, Disease Management, Epidemiology, Etiology, Prevention] | 1032 |
| 21 | underweight/di, dm, ep, et, pc [Diagnosis, Disease Management, Epidemiology, Etiology, Prevention] | 789 |
| 22 | stunting/di, dm, ep, et, pc [Diagnosis, Disease Management, Epidemiology, Etiology, Prevention] | 355 |
| 23 | chronic wasting disease/di, dm, ep, et, pc [Diagnosis, Disease Management, Epidemiology, Etiology, Prevention] | 234 |
| 24 | body mass index/ | 404144 |
| 25 | anthropometry/ | 47377 |
| 26 | (malnutrition or malnourish* or overnutrition or undernutrition or obese or obesity or overweight or adiposity or stunting or stunted or underweight or (wasting adj2 disease) or wasted or "body mass index" or BMI or anthropometr*).ti,ab,kw. | 859208 |
| 27 | or/15-26 | 961836 |
| 28 | 14 and 27 | 7182 |
| 29 | limit 28 to english | 6810 |

| | | |
|---|---|---|
| 30 | exp adolescence/ or exp adolescent/ or exp child/ or exp childhood disease/ or exp infant disease/ or (adolescen* or babies or baby or boy? or boyfriend or boyhood or girlfriend or girlhood or child* or girl? or infan* or juvenil* or juvenile* or kid? or minors or minors* or neonat* or neo-nat* or neo-nat* or newborn* or new-born* or paediatric* or peadiatric* or pediatric* or perinat* or preschool* or puber* or pubescen* or school or school child* or school* or schoolchild* or schoolchild* or pediatric* or paediatric* or infan* or child* or adolescen* or young).jn,jw. or (pediatric* or paediatric* or infan* or child* or adolescen* or young).in. or (teen* or toddler? or underage? or under-age? or youth*).ti,ab,kw. | 4255302 |
| 31 | 29 and 30 | 2812 |
| 32 | limit 31 to conference abstracts | 541 |
| 33 | limit 31 to (book or book series) | 10 |
| 34 | limit 31 to (books or chapter or conference abstract or letter or note or "review" or short survey) | 774 |
| 35 | limit 31 to (meta analysis or "systematic review") | 38 |
| 36 | 32 or 33 or 34 or 35 | 794 |
| 37 | 31 not 36 | 2018 |

**Table S3 Search strategy for Scopus.**

| # | Searching words | Results |
| --- | --- | --- |
| #1 | TITLE-ABS-KEY ( ( iron OR ferritin OR hemoglobin OR zinc OR "vitamin A" OR retinol ) AND ( serum OR blood OR plasma ) AND ( inadequa* OR insufficien* OR deficien* ) ) | 60369 |
| #2 | TITLE-ABS-KEY ( malnutrition OR malnourish* OR overnutrition OR undernutrition OR obese OR obesity OR overweight OR adiposity OR stunting OR stunted OR underweight OR ( wasting W/2 disease ) OR wasted OR "body mass index" OR bmi OR anthropometr* ) | 887265 |
| #3 | ( TITLE-ABS-KEY ( ( iron OR ferritin OR hemoglobin OR zinc OR "vitamin W/0 A" OR retinol ) AND ( serum OR blood OR plasma ) AND ( inadequa* OR insufficien* OR deficien* ) ) ) AND ( TITLE-ABS ( malnutrition OR malnourish* OR overnutrition OR undernutrition OR obese OR obesity OR overweight OR adiposity OR stunting OR stunted OR underweight OR ( wasting W/2 disease ) OR wasted OR "body mass index" OR bmi OR anthropometr* ) ) | 7,726 |
| #4 | ( TITLE-ABS-KEY ( ( iron OR ferritin OR hemoglobin OR zinc OR "vitamin W/0 A" OR retinol ) AND ( serum OR blood OR plasma ) AND ( inadequa* OR insufficien* OR deficien* ) ) ) AND ( TITLE-ABS-KEY ( malnutrition OR malnourish* OR overnutrition OR undernutrition OR obese OR obesity OR overweight OR adiposity OR stunting OR stunted OR underweight OR ( wasting W/2 disease ) OR wasted OR "body mass index" OR bmi OR anthropometr* ) ) AND ( LIMIT-TO ( LANGUAGE , "English" ) ) | 7,173 |
| #5 | ( TITLE-ABS-KEY ( ( iron OR ferritin OR hemoglobin OR zinc OR "vitamin W/0 A" OR retinol ) AND ( serum OR blood OR plasma ) AND ( inadequa* OR insufficien* OR deficien* ) ) ) AND ( TITLE-ABS-KEY ( malnutrition OR malnourish* OR overnutrition OR undernutrition OR obese OR obesity OR overweight OR adiposity OR stunting OR stunted OR underweight OR ( wasting W/2 disease ) OR wasted OR "body mass index" OR bmi OR anthropometr* ) ) AND ( ( TITLE-ABS-KEY ( adolescen* OR babies OR baby OR boy? OR boyfriend OR boyhood OR girlfriend OR girlhood OR child* OR girl? OR infan* OR juvenil* OR kid? OR minors OR minors* OR neonat* OR neo-nat* OR newborn* OR new-born* OR paediatric* OR peadiatric* OR pediatric* OR perinat* OR preschool* OR puber* OR pubescen* OR school* OR teen* OR toddler? OR underage? OR under-age? OR youth* ) ) ) AND ( LIMIT-TO ( LANGUAGE , "English" ) ) | 3,036 |
| #6 | ( TITLE-ABS-KEY ( ( iron OR ferritin OR hemoglobin OR zinc OR "vitamin W/0 A" OR retinol ) AND ( serum OR blood OR plasma ) AND ( inadequa* OR insufficien* OR deficien* ) ) ) AND ( TIT | 2,683 |

| | |
|---|---|
| | LE-ABS-KEY ( malnutrition OR malnourish* OR overnutrition OR undernutrition OR obese OR obesity OR overweight OR adiposity OR stunting OR stunted OR underweight OR ( wasting W/2 disease ) OR wasted OR "body mass index" OR bmi OR anthropometr* ) ) AND ( ( TITLE-ABS-KEY ( adolescen* OR babies OR baby OR boy? OR boyfriend OR boyhood OR girlfriend OR girlhood OR child* OR girl? OR infan* OR juvenil* OR kid? OR minors OR minors* OR neonat* OR neo-nat* OR newborn* OR new-born* OR paediatric* OR peadiatric* OR pediatric* OR perinat* OR preschool* OR puber* OR pubescen* OR school* OR teen* OR toddler? OR underage? OR under-age? OR youth* ) ) ) AND ( LIMIT-TO ( LANGUAGE , "English" ) ) AND ( LIMIT-TO ( DOCTYPE , "ar" ) ) |

**Table S4  Search strategy for Cochrane.**

| | | |
|---|---|---|
| #1 | MeSH descriptor: [Iron, Dietary] explode all trees and with qualifier(s): [blood - BL] | 54 |
| #2 | MeSH descriptor: [Iron] explode all trees and with qualifier(s): [blood - BL, deficiency - DF] | 933 |
| #3 | (((iron or ferritin or hemoglobin) and (serum or blood or plasma)) and (inadequa* or insufficien* or deficien*)):ti,ab,kw | 5477 |
| #4 | MeSH descriptor: [Anaemia, Iron-Deficiency] explode all trees | 1330 |
| #5 | #1 or #2 or #3 or #4 | 6310 |
| #6 | MeSH descriptor: [Zinc] explode all trees and with qualifier(s): [blood - BL, deficiency - DF] | 648 |
| #7 | (((zinc) and (serum or blood or plasma)) and (inadequa* or insufficien* or deficien*)):ti,ab,kw | 870 |
| #8 | #6 or #7 | 1243 |
| #9 | MeSH descriptor: [Vitamin A] tree(s) exploded and with qualifier(s): [blood - BL] | 497 |
| #10 | (((retinol or "vitamin A") and (serum or blood or plasma)) and (inadequa* or insufficien* or deficien*)):ti,ab,kw | 706 |
| #11 | #9 or #10 | 977 |
| #12 | #5 or #8 or #11 | 7724 |
| #13 | MeSH descriptor: [Obesity] explode all trees | 14007 |
| #14 | MeSH descriptor: [Thinness] explode all trees and with qualifier(s): [blood - BL, diagnosis - DI, etiology - ET, metabolism - ME, epidemiology - EP, prevention & control - PC] | 111 |
| #15 | MeSH descriptor: [Malnutrition] explode all trees and with qualifier(s): [blood - BL, diagnosis - DI, etiology - ET, metabolism - ME, epidemiology - EP, prevention & control - PC] | 1969 |
| #16 | MeSH descriptor: [Overweight] explode all trees and with qualifier(s): [blood - BL, diagnosis - DI, etiology - ET, metabolism - ME, epidemiology - EP, prevention & control - PC] | 6395 |
| #17 | MeSH descriptor: [Protein-Energy Malnutrition] explode all trees | 250 |
| #18 | MeSH descriptor: [Body Mass Index] explode all trees | 10188 |
| #19 | MeSH descriptor: [Anthropometry] explode all trees | 23419 |
| #20 | (malnutrition or malnourish* or overnutrition or undernutrition or obese or obesity or overweight or adiposity or stunting or stunted or underweight or (wasting NEAR/2 disease) or wasted or "body mass index" or BMI or anthropometr*):ti,ab,kw | 99958 |
| #21 | #13 or #14 or #15 or #16 or #17 or #18 or #19 or #20 | 109600 |
| #22 | #12 and #21 | 1831 |
| #23 | MeSH descriptor: [Adolescent] explode all trees | 104274 |
| #24 | MeSH descriptor: [Child] explode all trees | 56347 |
| #25 | MeSH descriptor: [Infant] explode all trees | 32245 |
| #26 | (infant disease* or childhood disease*):ti,ab,kw or (adolescen* or babies or baby or boy? or boyfriend or boyhood or girlfriend or girlhood or child* or girl? or infan* or juvenil* or kid? or minors or minors* or neonat* or neo-nat* or newborn* or new-born* | 308863 |

| | | |
|---|---|---|
| | or paediatric* or peadiatric* or pediatric* or perinat* or preschool* or puber* or pubescen* or school* or teen* or toddler? or underage? or under-age? or youth*):ti,ab,kw | |
| #27 | #23 or #24 or #25 or #26 | 308863 |
| #28 | #22 and #27 in trials | 882 |

**Table S5 Risk of bias assessment for included studies.**

| First Author, Year | Micronutrient | Nutrition | Q1 | Q2 | Q3 | Q4 | Q5 | Q6 | Q7 | Q8 | Q9 | Q10 | Overall Rating |
|---|---|---|---|---|---|---|---|---|---|---|---|---|---|
| Abd-El-Wahed, 2014 | Iron | Overnutrition | YES | YES | NO | NO | YES | YES | YES | NO | YES | NO | POSITIVE |
| Adamu, 2016 | VA | Undernutrition | YES | YES | NO | NO | YES | YES | YES | NO | NO | YES | POSITIVE |
| Ahmed, 2006 | VA | Undernutrition | YES | YES | YES | YES | YES | YES | YES | YES | YES | NO | POSITIVE |
| Alaofe, 2017 | Iron, VA | Undernutrition | YES | YES | YES | YES | YES | YES | YES | YES | YES | YES | POSITIVE |
| Al-Hussaini, 2022 | Iron, zinc | Undernutrition | YES | YES | YES | NO | YES | YES | NO | NO | YES | YES | NEUTRAL |
| Andre, 2017 | Iron | Undernutrition | YES | YES | YES | NO | YES | YES | YES | YES | NO | NO | POSITIVE |
| Brotanek, 2007 | Iron | Overnutrition | YES | YES | YES | NO | YES | YES | YES | YES | YES | YES | POSITIVE |
| Cabañas Pujadas, 2022 | Iron | Overnutrition | YES | YES | YES | NO | YES | YES | YES | YES | YES | YES | POSITIVE |
| Cepeda-Lopez, 2011 | Iron | Overnutrition | YES | YES | YES | NO | YES | YES | YES | YES | YES | YES | POSITIVE |
| Cheng, 2013 | Iron | Overnutrition | YES | YES | YES | NO | YES | YES | YES | YES | YES | YES | POSITIVE |
| Chitekwe, 2022 | Iron | Undernutrition | YES | YES | YES | YES | YES | YES | YES | YES | YES | YES | POSITIVE |
| Cobayashi, 2013 | Iron, VA | Both | YES | YES | YES | YES | YES | YES | YES | YES | YES | YES | POSITIVE |
| Dallazen, 2023 | VA | Both | YES | YES | YES | YES | YES | YES | YES | YES | YES | YES | POSITIVE |
| de Araújo, 2022 | Iron | Overnutrition | YES | YES | YES | YES | YES | YES | NO | NO | YES | YES | NEUTRAL |
| de Souza Valente da Silva, 2007 | VA | Overnutrition | YES | YES | YES | YES | YES | YES | YES | YES | NO | NO | POSITIVE |
| Dehghani, 2011 | Zinc | Both | YES | YES | NO | NO | YES | YES | YES | NO | NO | YES | POSITIVE |
| Disalvo, 2019 | VA | Both | YES | YES | YES | NO | YES | NO | YES | YES | YES | YES | POSITIVE |
| Eftekhari, 2009 | Iron | Both | YES | YES | NO | NO | YES | YES | YES | YES | NO | YES | POSITIVE |
| El Khoury, 2020 | Iron | Overnutrition | YES | YES | NO | NO | YES | YES | NO | NO | YES | NO | NEGATIVE |
| Engle-Stone, 2014 | Zinc | Undernutrition | YES | YES | YES | YES | YES | YES | YES | YES | NO | YES | POSITIVE |
| Ernawati, 2021 | Iron, VA | Undernutrition | YES | YES | YES | NO | YES | YES | YES | YES | YES | YES | POSITIVE |
| Fan, 2017 | Zinc | Overnutrition | YES | YES | YES | NO | YES | YES | YES | YES | YES | YES | POSITIVE |
| Ferrari, 2015 | Iron | Overnutrition | YES | YES | YES | NO | YES | YES | YES | YES | YES | YES | POSITIVE |
| Galetti, 2016 | Zinc | Undernutrition | YES | YES | YES | NO | YES | YES | YES | YES | NO | YES | NEUTRAL |
| Ghosh, 2012 | Iron | Undernutrition | YES | YES | NO | NO | YES | YES | NO | NO | NO | NO | NEGATIVE |
| Gibson, 2007 | Zinc | Undernutrition | YES | YES | YES | YES | YES | YES | YES | NO | NO | YES | POSITIVE |

| | | | | | | | | | | | | | |
|---|---|---|---|---|---|---|---|---|---|---|---|---|---|
| Goyena, 2021 | Zinc | Undernutrition | YES | YES | YES | YES | YES | YES | YES | YES | YES | YES | POSITIVE |
| Grant, 2007 | Iron | Overnutrition | YES | YES | YES | YES | YES | YES | YES | YES | NO | YES | POSITIVE |
| Gunanti, 2014 | VA | Overnutrition | YES | YES | YES | YES | YES | YES | YES | YES | YES | YES | POSITIVE |
| Habib, 2016 | Iron | Undernutrition | YES | YES | YES | YES | YES | YES | YES | YES | YES | YES | POSITIVE |
| Habib, 2022 | Zinc | Both | YES | YES | YES | NO | YES | YES | YES | NO | YES | YES | POSITIVE |
| Higgins, 2020 | Iron | Overnutrition | YES | YES | YES | NO | YES | YES | YES | YES | YES | YES | POSITIVE |
| Ho, 2017 | Zinc | Overnutrition | YES | Unclear | NO | NO | YES | YES | YES | NO | YES | YES | POSITIVE |
| Hu, 2001 | VA | Overnutrition | YES | YES | YES | NO | YES | YES | YES | YES | YES | NO | POSITIVE |
| Indriastuti Kurniawan, 2006 | Iron | Undernutrition | YES | YES | NO | NO | YES | YES | NO | NO | NO | NO | NEGATIVE |
| Kassem, 2022 | Iron | Overnutrition | YES | YES | YES | YES | YES | YES | YES | NO | YES | YES | POSITIVE |
| Khatib, 2009 | Iron, VA | Undernutrition | YES | YES | NO | YES | YES | YES | YES | NO | YES | YES | POSITIVE |
| Kongsbak, 2006 | Zinc | Undernutrition | YES | YES | YES | YES | YES | YES | YES | YES | YES | YES | POSITIVE |
| Kumari, 2022 | Iron, zinc | Undernutrition | YES | NO | YES | NO | YES | YES | YES | NO | YES | YES | POSITIVE |
| Kurugol, 2000 | VA | Undernutrition | YES | YES | NO | NO | YES | YES | YES | NO | NO | YES | NEUTRAL |
| Li, 2017 | Iron, zinc, VA | Both | YES | YES | YES | NO | YES | YES | YES | YES | YES | YES | POSITIVE |
| Lu, 2023 | Zinc | Both | YES | YES | YES | NO | YES | YES | YES | YES | YES | YES | POSITIVE |
| Manios,2013 | Iron | Overnutrition | YES | YES | YES | YES | YES | YES | YES | YES | YES | YES | POSITIVE |
| Marashinghe, 2015 | Zinc, VA | Undernutrition | YES | YES | YES | NO | YES | NO | YES | NO | YES | YES | NEUTRAL |
| Maslova, 2009 | Iron, VA | Both | YES | YES | YES | YES | YES | YES | YES | YES | YES | YES | POSITIVE |
| Matsungo, 2017 | Iron | Undernutrition | YES | YES | YES | NO | YES | YES | YES | YES | YES | YES | POSITIVE |
| Moschonis, 2012 | Iron | Overnutrition | YES | YES | YES | NO | YES | YES | NO | YES | YES | YES | POSITIVE |
| Nasiri-babadi, 2021 | Zinc | Undernutrition | YES | YES | YES | NO | YES | YES | YES | YES | YES | YES | POSITIVE |
| Naupal-Forcadilla, 2017 | Zinc | Undernutrition | YES | YES | YES | NO | YES | YES | YES | YES | NO | YES | POSITIVE |
| Nead, 2004 | Iron | Overnutrition | YES | YES | NO | NO | YES | YES | NO | YES | YES | NO | POSITIVE |
| Okafor, 2021 | Zinc | Undernutrition | YES | YES | YES | NO | YES | YES | YES | NO | YES | YES | NEUTRAL |
| Onabanjo, 2014 | Iron | Both | YES | NO | NO | YES | YES | YES | NO | NO | YES | YES | POSITIVE |
| Orsango, 2021 | Iron | Undernutrition | YES | YES | YES | NO | YES | YES | YES | YES | YES | YES | POSITIVE |
| Ortega-Senovilla, 2019 | VA | Overnutrition | YES | YES | YES | NO | YES | YES | NO | NO | YES | YES | NEUTRAL |
| Ortiz Perez, 2020 | Iron | Overnutrition | YES | YES | NO | YES | YES | YES | YES | NO | YES | YES | POSITIVE |

| | | | | | | | | | | | | | |
|---|---|---|---|---|---|---|---|---|---|---|---|---|---|
| Oso, 2003 | VA | Undernutrition | YES | YES | NO | YES | YES | YES | YES | NO | NO | NO | NEUTRAL |
| Paes-Silva, 2018 | VA | Overnutrition | YES | YES | NO | NO | YES | YES | YES | YES | YES | YES | POSITIVE |
| Park, 2012 | Iron, zinc | Undernutrition | YES | YES | YES | NO | YES | YES | YES | YES | YES | YES | POSITIVE |
| Perng, 2013 | Iron | Both | YES | YES | YES | NO | YES | YES | YES | NO | YES | YES | POSITIVE |
| Perrone, 1998 | Zinc | Overnutrition | YES | NO | NO | NO | YES | YES | YES | NO | YES | NO | NEUTRAL |
| Pompano, 2022 | Iron | Overnutrition | YES | YES | YES | YES | YES | YES | YES | NO | YES | YES | POSITIVE |
| Samba, 2006 | VA | Undernutrition | YES | YES | NO | NO | YES | YES | YES | NO | NO | YES | POSITIVE |
| Sethy, 2014 | Iron, zinc | Undernutrition | YES | NO | NO | NO | YES | YES | YES | NO | NO | NO | NEGATIVE |
| Sharif, 2019 | zinc, VA | Overnutrition | YES | YES | YES | YES | YES | YES | YES | YES | YES | YES | POSITIVE |
| Shattnawi, 2018 | Iron | Overnutrition | YES | YES | YES | YES | YES | YES | YES | YES | YES | YES | POSITIVE |
| Shukla, 2023 | Iron | Undernutrition | YES | NO | YES | NO | YES | YES | NO | NO | NO | YES | NEUTRAL |
| Ssentongo, 2020 | VA | Undernutrition | YES | YES | YES | NO | YES | YES | YES | YES | YES | YES | POSITIVE |
| Suteerojntrakoo, 2021 | Iron | Overnutrition | YES | YES | YES | YES | YES | YES | NO | YES | YES | YES | NEUTRAL |
| Sypes, 2018 | Iron | Overnutrition | YES | YES | YES | YES | YES | YES | YES | YES | YES | YES | POSITIVE |
| Tan, 2023 | Iron | Both | YES | YES | YES | NO | YES | YES | YES | YES | YES | YES | POSITIVE |
| Tessema, 2019 | Zinc | Undernutrition | YES | YES | NO | YES | YES | NO | YES | YES | YES | YES | POSITIVE |
| Thillan, 2021 | Iron, zinc, VA | Overnutrition | YES | YES | YES | NO | YES | YES | YES | NO | YES | YES | POSITIVE |
| Tian, 2022 | VA | Overnutrition | YES | YES | YES | NO | YES | YES | YES | YES | YES | YES | POSITIVE |
| Tussing-Humphreys, 2009 | Iron | Overnutrition | YES | YES | YES | NO | YES | YES | YES | YES | YES | YES | POSITIVE |
| Van Nhien, 2009 | Zinc | Undernutrition | YES | YES | NO | YES | YES | YES | YES | NO | YES | YES | POSITIVE |
| Wei, 2016 | VA | Overnutrition | YES | YES | YES | YES | YES | YES | YES | YES | YES | YES | POSITIVE |
| Yalcin, 2019 | Iron, zinc | Overnutrition | YES | YES | YES | NO | YES | YES | YES | NO | YES | YES | POSITIVE |
| Yang, 2015 | VA | Overnutrition | YES | YES | YES | NO | YES | YES | YES | NO | YES | YES | POSITIVE |
| Yazbeck, 2016 | Zinc | Undernutrition | YES | YES | NO | NO | YES | YES | YES | NO | YES | YES | POSITIVE |
| Zhu, 2019 | Iron | Both | YES | YES | YES | NO | YES | YES | NO | YES | YES | UES | POSITIVE |
| Zhu, 2021 | Zinc | Overnutrition | YES | YES | YES | YES | YES | YES | YES | YES | YES | YES | POSITIVE |
| Zimmermann, 2008 | Iron | Both | YES | Unclear | YES | NO | YES | YES | YES | YES | YES | YES | POSITIVE |
| Zou, 2022 | Zinc, VA | Overnutrition | YES | YES | YES | NO | YES | YES | YES | YES | YES | YES | POSITIVE |

**Figure S1** Subgroup analysis of associations between iron deficiency and overnutrition stratified by gender. The vertical line represented no effect (OR=1.00) and the overall effect (OR=1.51).

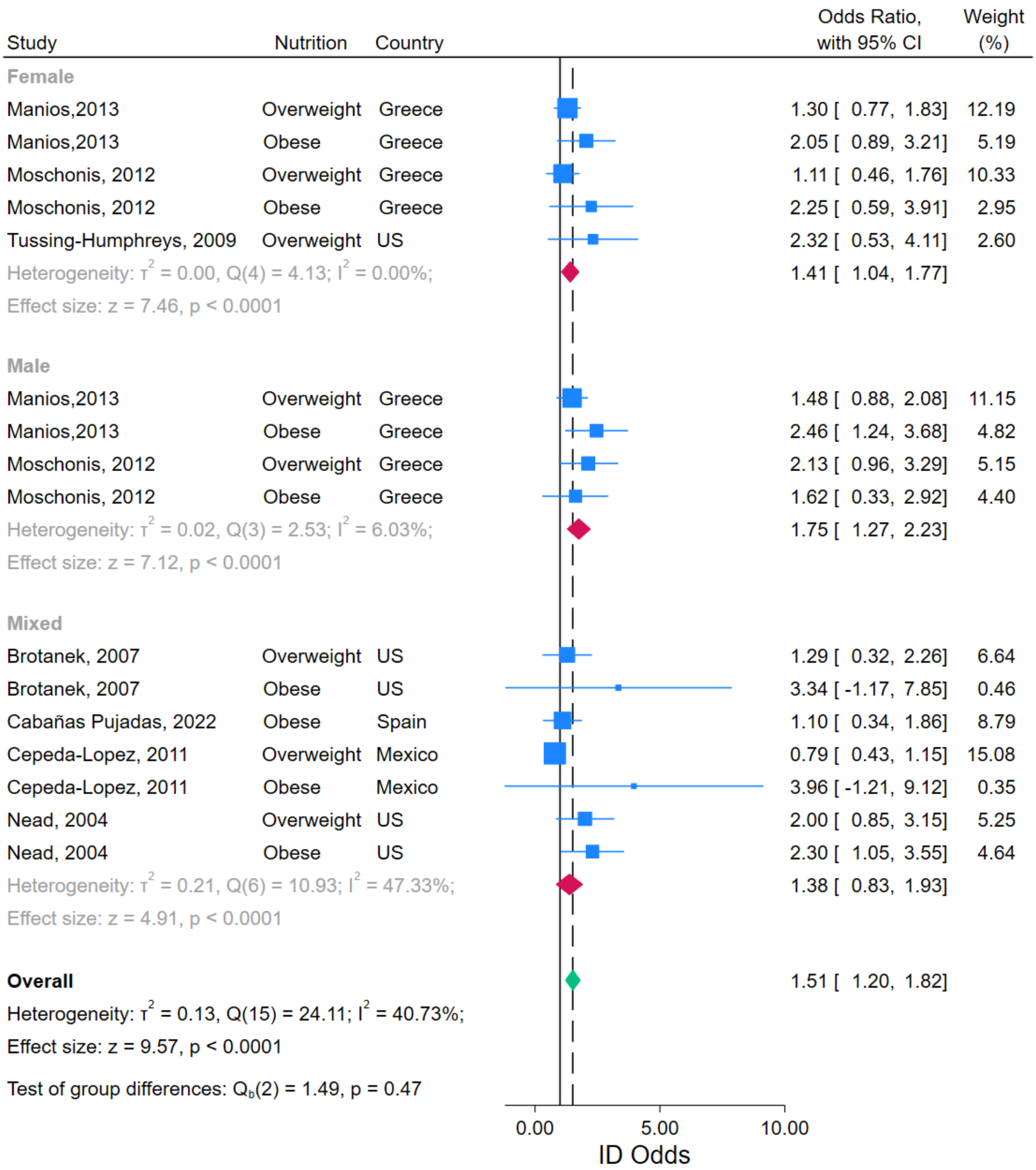

**Figure S2** Funnel plots for nutrition status and gender. **A.** nutrition status and **B.** gender. The red line represents the odds ratio for each group.

**A.**

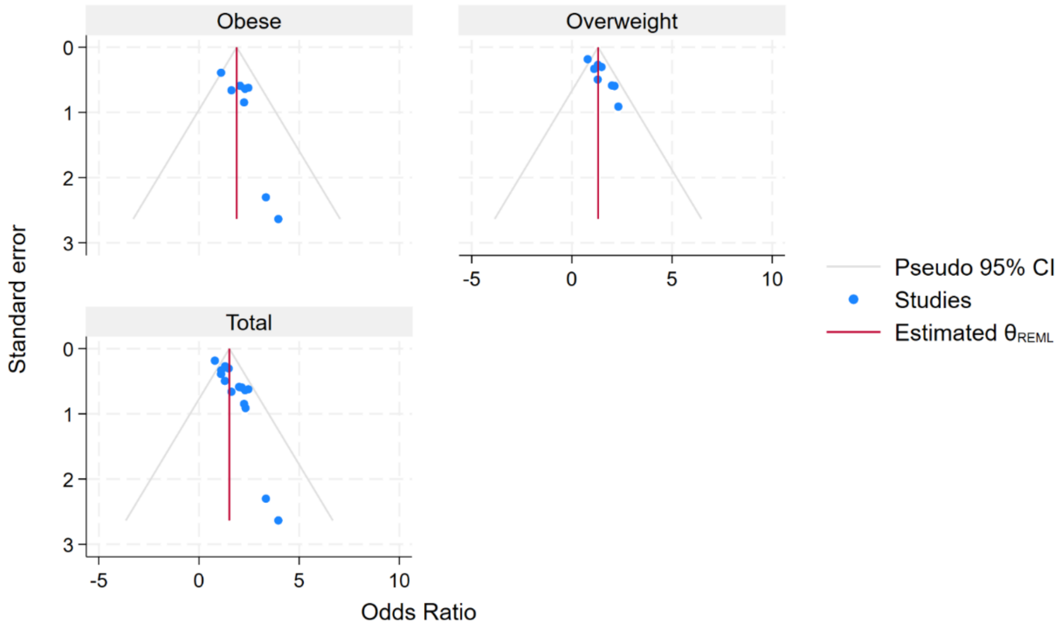

**B.**

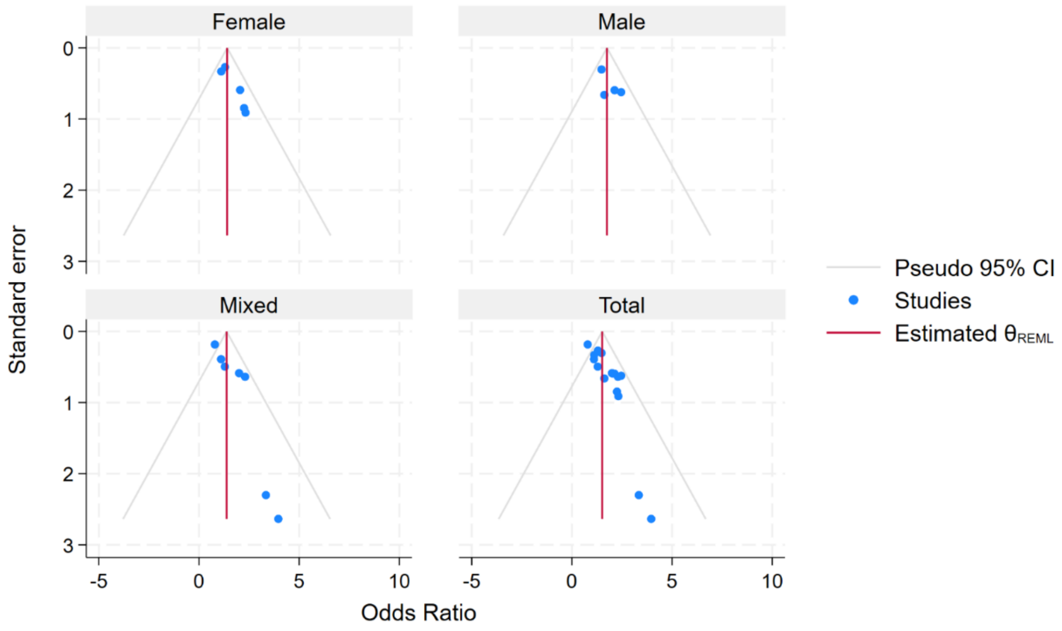

**Figure S3** Leave-one-out sensitivity test for overall and subgroup effect size. The red line represented the effect size before omitting any study. **A.** Overall **B.** obese, **C.** overweight.

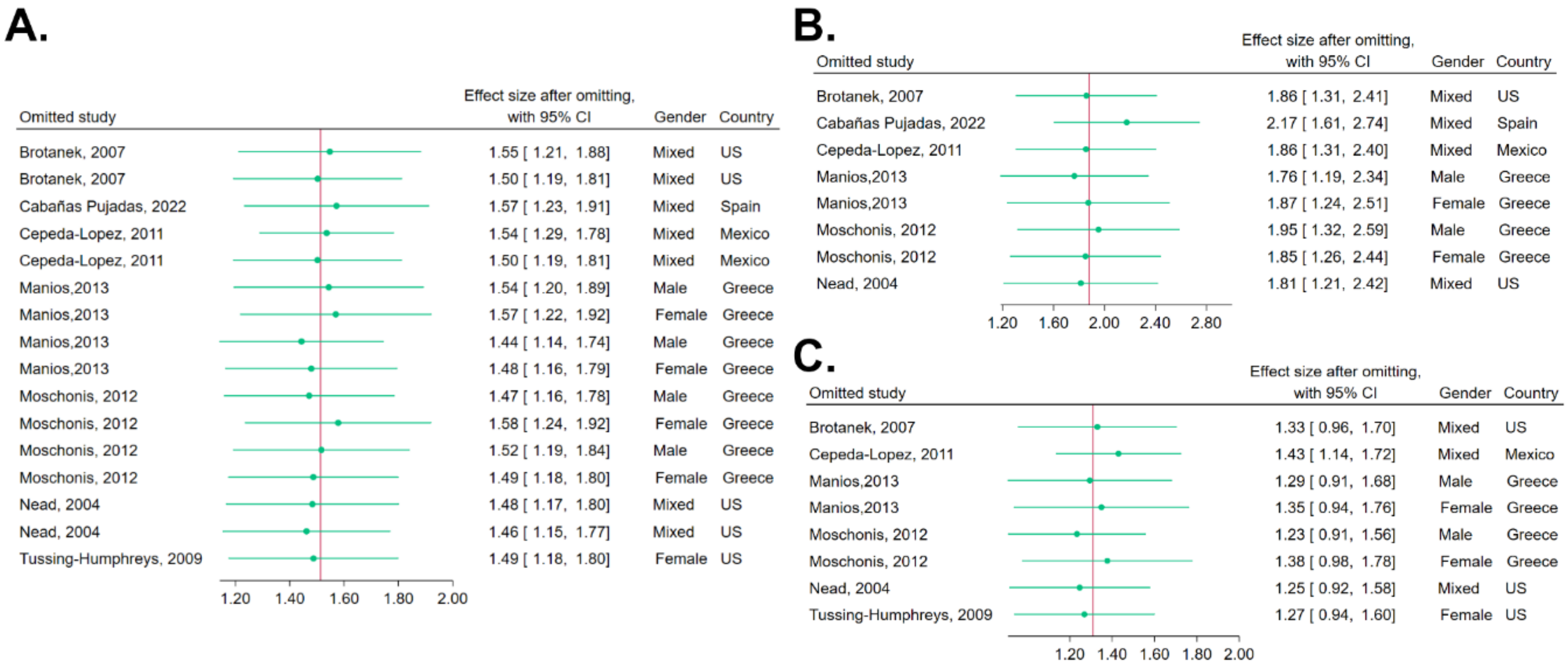
